# Plasma protein biomarkers distinguish Multisystem Inflammatory Syndrome in Children (MIS-C) from other pediatric infectious and inflammatory diseases

**DOI:** 10.1101/2023.07.28.23293197

**Authors:** Sophya Yeoh, Diego Estrada-Rivadeneyra, Heather Jackson, Ilana Keren, Rachel Galassini, Samantha Cooray, Priyen Shah, Philipp Agyeman, Romain Basmaci, Enitan Carrol, Marieke Emonts, Colin Fink, Taco Kuijpers, Federico Martinon- Torres, Marine Mommert-Tripon, Stephane Paulus, Marko Pokorn, Pablo Rojo, Lorenza Romani, Luregn Schlapbach, Nina Schweintzger, Ching-Fen Shen, Maria Tsolia, Effua Usuf, Michiel Van der Flier, Clementien Vermont, Ulrich Von Both, Shunmay Yeung, Dace Zavadska, Lachlan Coin, Aubrey Cunnington, Jethro Herberg, Michael Levin, Myrsini Kaforou, Shea Hamilton, the PERFORM, DIAMONDS, UK KD Genetics Consortia

## Abstract

**Background:** Multisystem inflammatory syndrome in children (MIS-C) is a rare but serious hyperinflammatory complication following infection with severe acute respiratory syndrome coronavirus 2 (SARS-CoV-2). The mechanisms underpinning the pathophysiology of MIS-C are poorly understood. Moreover, clinically distinguishing MIS-C from other childhood infectious and inflammatory conditions, such as Kawasaki Disease (KD) or severe bacterial and viral infections is challenging due to overlapping clinical and laboratory features. We aimed to determine a set of plasma protein biomarkers that could discriminate MIS-C from those other diseases.

**Methods:** Seven candidate protein biomarkers for MIS-C were selected based on literature and from whole blood RNA-Sequencing data from patients with MIS-C and other diseases. Plasma concentrations of ARG1, CCL20, CD163, CORIN, CXCL9, PCSK9 and ADAMTS2 were quantified in MIS-C (n=22), KD (n=23), definite bacterial (DB; n=28) and viral (DV, n=27) disease, and healthy controls (n=8). Logistic regression models were used to determine the discriminatory ability of individual proteins and protein combinations to identify MIS-C, and association with severity of illness.

**Results:** Plasma levels of CD163, CXCL9, and PCSK9 were significantly elevated in MIS-C with a combined AUC of 86% (95% CI: 76.8%-95.1%) for discriminating MIS-C from other childhood diseases. Lower ARG1 and CORIN plasma levels were significantly associated with severe MIS-C cases requiring oxygen, inotropes or with shock.

**Conclusion:** Our findings demonstrate the feasibility of a host protein biomarker signature for MIS-C and may provide new insight into its pathophysiology.

## INTRODUCTION

Although children have been generally less severely affected than adults by COVID-19 infection, a small proportion of children develop a rare but serious hyperinflammatory condition termed Multisystem Inflammatory Condition in Children (MIS-C) or Pediatric Inflammatory Multisystem Syndrome temporally associated with COVID-19 (PIMS-TS) (1) MIS-C usually develops 2-6 weeks following SARS-CoV-2 infection, with children presenting with febrile illness and a multisystem hyperinflammatory state that shows some overlapping clinical characteristics with other infectious and inflammatory childhood disorders including Kawasaki Disease (KD) and severe bacterial infections such as Toxic Shock Syndrome (TSS) (2,3,4).

MIS-C leads to critical illness in ∼70% of affected children and, as of July 2023, the CDC have reported 9,499 cases meeting the full clinical definitions of this novel disorder (5). MIS-C has been reported to have a higher incidence in older children, males, and in children of Black, Asian, or Hispanic ethnicity (6,7,8). Common symptoms of MIS-C include persistent fever, oral mucosal inflammation, conjunctivitis, skin rash, elevated inflammatory markers, gastrointestinal involvement, cardiac manifestations, multisystem organ dysfunction, and shock (9).

KD is an acute systemic vascular disease of unknown etiology affecting predominately the coronary arteries in infants and children (10,11). The standard treatment for KD patients is high-dose intravenous immunoglobulin (IVIG) which can quickly alleviate symptoms and reduce the incidence of coronary artery aneurysms (12). As there is currently no diagnostic test for KD, the diagnosis heavily relies on clinical signs and symptoms which include high levels of markers of inflammation and mucosal changes. Approximately 40% of patients with MIS-C will meet the diagnostic criteria for KD and due to the overlapping clinical and laboratory features, IVIG was recommended as a first-line treatment for MIS-C (5,13). However, recent evidence has shown that treatment of MIS-C with corticosteroids has the same outcome as IVIG treatment, which is important because corticosteroids are cheap and readily available in low– and middle-income countries (14,15). A diagnostic test to distinguish between these two diseases would therefore greatly benefit treatment and patient outcome.

Host-derived protein biomarkers have been successfully used to distinguish bacterial from viral infection in children (16). It has been reported that elevated levels of soluble biomarkers associated with inflammation, vascular endothelial injury, mucosal immune dysregulation, septic shock, and cardiac and GI involvement have been observed in MIS-C (2,6,7,8,17,18,19,20). From a proteomic perspective, it has also been suggested that there are clinical similarities between MIS-C and secondary causes of Hemophagocytic Lymphohistiocytosis such as Macrophage Activation Syndrome (HLH/MAS) and Thrombotic Microangiopathy (TMA), in addition to its clinical similarities with KD and TSS (19). These studies have singled out potential individual biomarkers of MIS-C, such as Cysteine–Cysteine Motif Chemokine Ligand 20 (CCL20), C-X-C Motif Chemokine Ligands 9 and 10 (CXCL9, CXCL10), Interferon-gamma (IFN-g), Interleukins 6, 7, 8 and 10 (IL-6, IL-7, IL-8 and IL-10), Phospholipase A2 Group IIA (PLA2G2A), Tumor Necrosis Factor alpha (TNF-a), Nuclear Factor Kappa B (NF-κB) and Zonulin. However, the results differ between studies, which may be due to the different comparator groups that were used.

In this study, we aimed to identify a set of proteins that could distinguish MIS-C from other disease groups, including KD, definite bacterial infections (DB), and definite viral infections (DV) by quantifying plasma protein levels of candidate MIS-C biomarkers. Such a protein signature could be used as the basis for a diagnostic test. We also aimed to associate significant changes in individual protein levels with clinical presentation, such association could be further investigated to predict disease severity or inform clinical management.

## MATERIALS AND METHODS

### Ethics Statement

Informed consent was taken from all patients recruited to the study. Recruitment at all locations took place after ethical permissions were in place for that area. In the DIAMONDS study (Diagnosis and Management of Febrile Illness using RNA Personalized Molecular Signature Diagnosis https://www.diamonds2020.eu) and the PERFORM study (Personalized Risk assessment in Febrile illness to Optimize Real-life Management, perform2020.org/), the consortium agreed on a finalized protocol and supporting documents that were translated into local languages. Each participating country took responsibility for gaining ethical approval in their region.

For DIAMONDS, the lead site received ethical approval for United Kingdom centers from the London-Dulwich research ethics committee (20/HRA/1714). For the PERFORM study, the lead site received approval ethical approval for United Kingdom centers from the London-Central research ethics committee (16/LO/1684). The Genetic Determinants of Kawasaki Disease for Susceptibility and Outcome study recruited in the United Kingdom only, and ethical approval was granted by the London-Fulham research ethics committee (13/LO/0026). During the conduct of the study, clinical data and samples were identified only by anonymized study numbers.

### Study Population

This study included 108 pediatric patients (≤16 years old) with clinical and laboratory-confirmed MIS-C (n = 22), Kawasaki disease (KD, n = 23), definite bacterial infections (DB, n = 28), definite viral infections (DV, n = 27), and healthy controls (HC, n = 8). A breakdown of the definite bacterial and viral infections can be found in Figure S1.

The WHO defines MIS-C as children 0-19 years of age with fever ≥ 3 days and elevated inflammatory markers (including C-reactive protein, procalcitonin, and erythrocyte sedimentation rate) who have no other obvious microbial cause of inflammation, including bacterial sepsis, staphylococcal or streptococcal shock syndromes and evidence of SARS-CoV-2 infection or probable exposure (21). They must also exhibit two of the most common features of MIS-C including gastrointestinal (GI) symptoms, rash or bilateral conjunctivitis, hypotension, shock, or cardiac complications. All of the MIS-C patients met the WHO criteria, which was the recommended criteria at the time.

Plasma samples from all of the other disease groups (KD, DB, DV) were collected prior to the COVID-19 pandemic (October 2013-January 2019). Plasma samples from KD patients were collected prior to receiving IVIG treatment in 21 out of 23 cases used in this study. All DB and DV patients were phenotyped according to our published algorithm and as previously described (22,23,24). In brief, the DB group included patients in whom an appropriate bacterial pathogen was isolated from a normally sterile site and a diagnosis of DV was conditional upon identification of a virus compatible with the clinical syndrome with no evidence of bacterial infection and CRP ≤ 60mg/L. No deaths occurred in this study population.

### Protein Selection

Proteins were selected either through candidate gene biomarkers identified by RNA Sequencing (RNA-Seq) or through existing literature at the time of this study design.

Whole blood transcriptome profiling was performed using RNA-Seq on samples obtained from patients with MIS-C, KD, DB infections, DV infections, and HC children. Following pre-processing and normalization which is described in detail in Jackson et al., (25), differential expression analysis was performed to identify the genes that were significantly differentially expressed (SDE) between MIS-C and KD, DB infections, and DV infections. Differential expression analysis was performed using DESeq2 with models including age, sex at birth, and RNA-Seq batch, as two experimental runs were used for sequencing (26). The following comparisons were made: MIS-C vs. KD+DB+DV; MIS-C vs. KD; MIS-C vs. DB; and MIS-C vs. DV. P-values were adjusted for multiple testing using the Benjamini-Hochberg adjustment and genes with adjusted p-values <0.05 were considered SDE (27).

Genes of interest were selected based on the criteria that they were significantly up-regulated in MIS-C cases as compared to one or more of the other disease groups. Due to the high number of SDE genes, we selected genes with BH-adjusted p-values < 10^-5, and a log-fold change >2, and those that encoded for extracellularly secreted proteins for which antibodies were commercially available were considered. The final proteins chosen based on the RNA-Seq data were ADAMTS2, ARG1, CD163, CORIN, and PCSK9. Two additional protein targets, CCL20 and CXCL9, were selected based on existing MIS-C literature at the time of this study because of their reported roles in the gastrointestinal involvement and hyper-inflammatory state observed early in the course of MIS-C disease, respectively (6,17,28).

### Enzyme-Linked Immunosorbent Assay (ELISA)

Plasma concentrations of ADAMTS2 were quantified using a commercial ELISA kit, Human ADAMTS2 (Abexxa, abx519366) according to the manufacturer’s protocol. Samples were randomized, diluted 1:10 in sample diluent buffer, and run in duplicate. Absorbance was measured at 450 nm on a SpectraMax microplate reader (Molecular Devices) and a standard curve was created using the SoftMax Pro software (version 5.0). Samples that fell below the lower limit of quantification (LLOQ) were reanalyzed and values that were still below the LLOQ were extrapolated as half of the lowest standard value.

### Multiplex Immunoassay

Plasma concentrations of ARG1, CCL20, CD163, CORIN, CXCL9, and PCSK9 were quantified using customized multiplex immunoassay kits from MSD (Meso Scale Discovery, Rockville, MD). CD163 (MSD, F21J4) and PCSK9 (MSD, F21ABA) were multiplexed on a 2-assay plate (MSD, K15227N) according to the manufacturer’s instructions with samples diluted 1:20 in assay diluent and run in duplicate. ARG1 (MSD, F21Q1), CCL20 (MSD, B21UZ), CORIN (MSD, F210B), and CXCL9 (MSD, F210I) were multiplexed on a 4-assay plate (MSD, K15229N) according to the manufacturer’s instructions with samples diluted 1:2 in assay diluent and run in duplicate. Two control plasma samples were included on each plate to account for inter-plate variation. All plates were analyzed on the Meso Quick Plex SQ120 Instrument (MSD). Data was generated on the Methodological Mind software (version 1.0.36) and analyzed using Discovery Workbench software (version 4.0, MSD).

### Statistical Analysis

All statistical analyses were performed using R (version 4.1.1) (29). Individual protein results were analyzed using one-way analysis of variance (ANOVA), followed by Tukey’s honestly significant difference (HSD) test to evaluate the differences between each cohort. Principal component analysis (PCA) was used to visualize the data. ANOVA p-values were corrected using the Bonferroni procedure and Tukey HSD test p-values were corrected using the in-build correction as part of the Tukey HSD test.

Associations between each of the protein markers and clinical variables were tested using unpaired two-tailed t-tests for categorical variables and linear regression models for continuous variables with all p-values corrected for multiple testing using the Bonferroni procedure (corrected per clinical/laboratory variable). Clinical and laboratory variables included admission to the pediatric intensive care unit (PICU), oxygen or ventilation requirement (invasive or non-invasive), inotrope administration, duration of symptoms at the time samples were taken, neutrophils, lymphocytes, platelets, CRP, white blood cell (WBC) counts, and presentation of symptoms including cardiac and gastrointestinal involvement, rash, shock, and conjunctivitis.

The distinguishing ability of each of the proteins that were significantly differentially abundant between MIS-C and other disease groups was evaluated using the area under the receiver operating characteristic (ROC) curve (AUC).

The performance of the proteins when combined into a multivariate model was also evaluated, with the proteins combined using the simple disease risk score (DRS) first described in Herberg et al., (22) and calculated as follows:

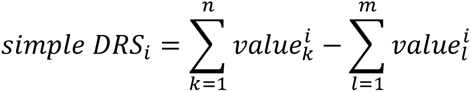

where *n* and *m* are the proteins that increase and decrease, respectively, in MIS-C *vs*. other disease groups. CRP was added to the protein model to determine whether its inclusion would improve model performance.

## RESULTS

### Clinical Features

108 samples were included in this study, with the patient demographic and clinical characteristics summarized in Table 1. All 22 MIS-C patients met the WHO case definitions for inclusion (21). A higher proportion of cases were male (68.2%, n=15) and the median age of the patients was higher in MIS-C than in the other disease comparator groups. Over half of MIS-C cases (n=12, 54.5%) required PICU admission and the majority of cases (n=18, 81.8%) presented with gastrointestinal symptoms.

**Table 1.** Demographic and clinical characteristics of the patients with MIS-C, DB, DV, and KD used for the ELISA and Meso Scale Discovery validation assays. Values are median (IQR) unless stated.

|  | MIS-C (n=22) | DB (n=27) | DV (n=28) | KD (n=23) | HC (n=8) |
| --- | --- | --- | --- | --- | --- |
| **Demographics** |  |  |  |  |  |
| **Age** | 8.0 (6.0 - 12.2) | 3.8 (2.4 - 8.3) | 1.1 (0.4 - 3.4) | 3.0 (1.4 - 4.0) | 12.8 (7.3 - 14.8) |
| **Sex (male, %)** | 15 (68%) | 15 (56%) | 14 (50%) | 14 (61%) | 6 (75%) |
| **Race (n, %)** |  |  |  |  |  |
| White/European | 10 (45%) | 22 (82%) | 14 (50%) | 7 (30%) | 8 (100%) |
| East/West/ South Asian | 6 (27%) | 2 (7%) | 8 (29%) | 6 (26%) | 0 (0%) |
| Black/African/ Caribbean | 3 (14%) | 1 (4%) | 4 (14%) | 7 (30%) | 0 (0%) |
| Other | 3 (14%) | 2 (7%) | 2 (7%) | 3 (13 %) | 0 (0%) |
| **Sample period** | Aug 2020 - Sep 2021 | Dec 2016 - Jan 2019 | Feb 2017 - Jan 2019 | Oct 2013 - Jun 2018 | Aug 2018 - Mar 2020 |
| **Treatment** |  |  |  |  |  |
| **Antibiotics (n, %)** | 22 (100%) | 18 (67%) | 14 (50%) | 23 (100%) | 0 (0%) |
| **Inotropes (n, %)** | 6 (27%) | 4 (15%) | 3 (11%) | 0 (0%) | 0 (0%) |
| **Oxygen (n, %)** | 3 (14%) | 9 (33%) | 14 (50%) | 0 (0%) | 0 (0%) |
| **Non-invasive ventilation (n, %)** | 2 (9%) | 1 (4%) | 10 (36%) | 0 (0%) | 0 (0%) |
| **Invasive ventilation (n, %)** | 1 (5%) | 7 (26%) | 14 (50%) | 0 (0%) | 0 (0%) |
| **Clinical features** |  |  |  |  |  |
| **Days from onset of symptoms** | 6.5 (5.0 - 7.0) | 3.0 (2.0 - 5.5) | 3.0 (2.0 - 5.0) | 6.0 (5.0 - 8.0) | N/A |
| **Admitted to PICU (n, %)** | 12 (55%) | 10 (37%) | 16 (57%) | 1 (4%) | 0 (0%) |
| **GI involvement (n, %)** | 18 (82%) | 3 (11%) | 2 (7%) | 4 (17%) | 1 (13%) |
| **Cardiac (n, %)** | 3 (14%) | 0 | 1 (4%) | 9 (39%) | 0 (0%) |
| **Pulmonary (n, %)** | 1 (5%) | 2 (7%) | 5 (18%) | N/A | 0 (0%) |
| **Neurological (n, %)** | 3 (14%) | 2 (7%) | 4 (14%) | N/A | 0 (0%) |
| **Shock (n, %)** | 5 (23%) | 5 (19%) | 2 (7%) | N/A | 0 (0%) |
| **Sepsis (n, %)** | 0 | 13 (48%) | 5 (18%) | N/A | 0 (0%) |
| **Comorbidities (n, %)** | 8 (36%) | 7 (26%) | 11 (39%) | 13 (57%) | 0 (0%) |
| **Blood parameters** |  |  |  |  |  |
| **Max CRP (mg/L)** | 241.0 (191.0 - 301.8) | 225.0 (125.7 - 343.6) | 19.2 (4.9 - 35.2) | 89.1 (62.8 - 136.0) | N/A |
| **Haemoglobin (g/L)** | 107.0 (95.8 - 118.8) | 104.0 (94.3 - 115.8) | 104.0 (91.0 - 123.8) | N/A | N/A |
| **Creatinine (µmol/L)** | 44.5 (35.3 - 52.8) | 44.1 (27.1 - 59.7) | 33.0 (28.0 - 47.0) | N/A | N/A |
| **ALT (IU/L)** | 25.5 (21.0 - 36.0) | 11.0 (10.0 - 23.0) | 24.0 (16.5 - 38.5) | N/A | N/A |
| **Bilirubin (µmol/L)** | 5.0 (5.0 - 7.0) | 8.5 (4.3 - 16.5) | 5.0 (3.5 - 12.3) | N/A | N/A |
| **Albumin (g/L)** | 24.5 (22.3 - 27.0) | 32.5 (29.3 - 37.3) | 30.5 (26.0 - 34.8) | N/A | N/A |
| **White blood cells (10<sup>9</sup>/L)** | 11.5 (6.0 - 16.3) | 17.4 (13.6 - 27.1) | 7.6 (5.7 - 10.8) | 12.3 (10.6 - 14.1) | N/A |
| **Neutrophils (10<sup>9</sup>/L)** | 8.4 (5.9 - 12.2) | 13.3 (9.7 - 19.4) | 3.3 (1.6 - 4.7) | 8.3 (6.9 - 11.2) | N/A |
| **Lymphocytes (10<sup>9</sup>/L)** | 1.3 (0.6 - 1.8) | 2.5 (1.8 - 3.7) | 3.1 (2.1 - 4.5) | 2.5 (1.7 - 3.0) | N/A |
| **Monocytes (10<sup>9</sup>/L)** | 0.4 (0.2 - 0.8) | 1.2 (0.9 - 2.3) | 0.8 (0.6 - 1.3) | 0.7 (0.5 - 0.9) | N/A |
| **Platelets (10<sup>9</sup>/L)** | 238.5 (113.3 - 321.8) | 269.5 (221.0 - 430.0) | 272.0 (230.0 - 321.0) | 330.0 (62.8 - 136.0) | N/A |

IQR: Interquartile range; N/A: Not available/Not applicable; MIS-C: Multisystem inflammatory syndrome in children; KD: Kawasaki disease; HC: Healthy controls; PICU: Paediatric intensive care unit; GI: Gastrointestinal; ALT: Alanine aminotransferase; CRP: C-reactive protein.

### Candidate Protein Biomarkers

Seven protein biomarkers for distinguishing MIS-C from other pediatric infectious and inflammatory diseases were selected through unbiased analyses of host transcriptomic profiles obtained from children with MIS-C (25) as well as from the literature. These included ADAMTS2, ARG1, CD163, CORIN, and PCSK9 which were selected from RNA-Seq, and CCL20 and CXCL9 which were selected from the literature (Table S1). Genes were selected based on BH-adjusted p-values < 10^-5, a log-fold change >2, and if they encoded for extracellularly secreted proteins.

All proteins were measured in a cohort of children with MIS-C, KD, DB or DV infections, and HC. When visualized using principal component analysis (PCA), the disease group was clearly captured by principal component (PC) 1, with a correlation of 47% (p-value: 3.3×10^-7^) between PC1 and whether a patient had MIS-C. Sex and age had a correlation of –1.8% and 7.9%, respectively, with PC1 (Figure S2).

### CD163, CXCL9, and PCSK9 are Significantly Differentially Abundant (SDA) in MIS-C *vs*. other diseases and healthy controls

Pediatric plasma samples were analyzed from MIS-C (n = 22), KD (n = 23), DB (n = 28), DV (n = 27), and healthy controls (n = 8). The protein concentrations were compared between MIS-C and other diseases (KD, DB, and DV combined) and healthy controls to evaluate how well the proteins would be able to discriminate MIS-C from non-MIS-C groups (e.g., MIS-C *vs*. KD, DB, DV, HC). The protein concentrations were then compared between MIS-C against each diagnostic group individually. When protein abundance levels were compared between MIS-C and all other comparator groups combined (i.e., HC, KD, DB, DV), significant differences were observed for PCSK9 (Bonferroni adjusted p-value: 3.3×10^-5^; Figure 1A), CD163 (Bonferroni adjusted p-value: 2.9×10^-4^; Figure 1B), and CXCL9 (Bonferroni adjusted p-value: 2.4×10^-2^; Figure 1C). For CD163, CXCL9, and PCSK9, levels were elevated in MIS-C *vs*. all other comparator groups, with significant *p*-values when pairwise comparisons were performed using the Tukey test, for all comparisons except for MIS-C *vs*. DV for CXCL9 (Table S2). ADAMTS2, ARG1, CCL20, and CORIN showed no significant difference (adjusted p-value >0.05) between MIS-C and both healthy controls and other diseases, nor between MIS-C and each disease group individually.

**Figure 1.**
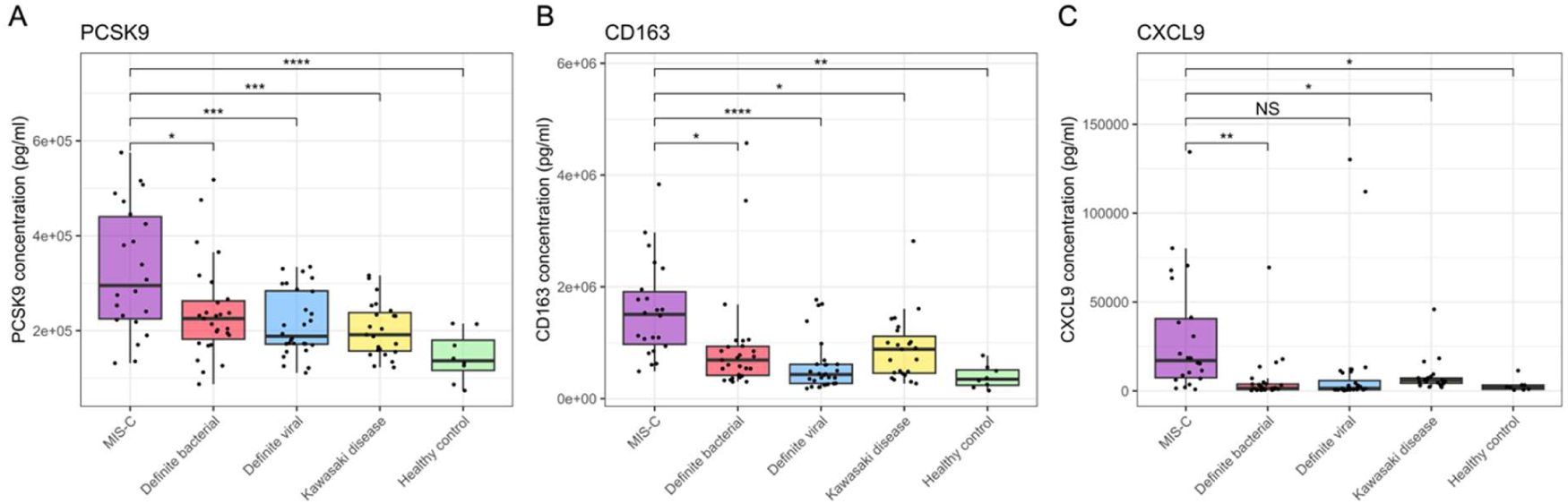
Boxplots showing concentrations of proteins with significantly different levels between MIS-C and the comparator groups (KD, DB, DV, HC). Boxplots are shown for PCSK9 (A), CD163 (B) and CXCL9 (C). MIS-C = multisystem inflammatory syndrome in children; KD = Kawasaki disease; DB = definite bacterial; DV = definite viral; HC = healthy control. ns= P > 0.05; * = P ≤ 0.05; ** = P ≤ 0.01, *** = P ≤ 0.001; **** = P ≤ 0.0001.

### A 3-protein signature can distinguish MIS-C from other pediatric infectious and inflammatory diseases

The performance of PCSK9, CD163, and CXCL9 when combined into a 3-protein signature was evaluated, returning an overall AUC of 86.09% (95% CI: 76.8%-95.1%) for distinguishing between MIS-C and DB, DV, and KD (Figure 2A).

**Figure 2.**
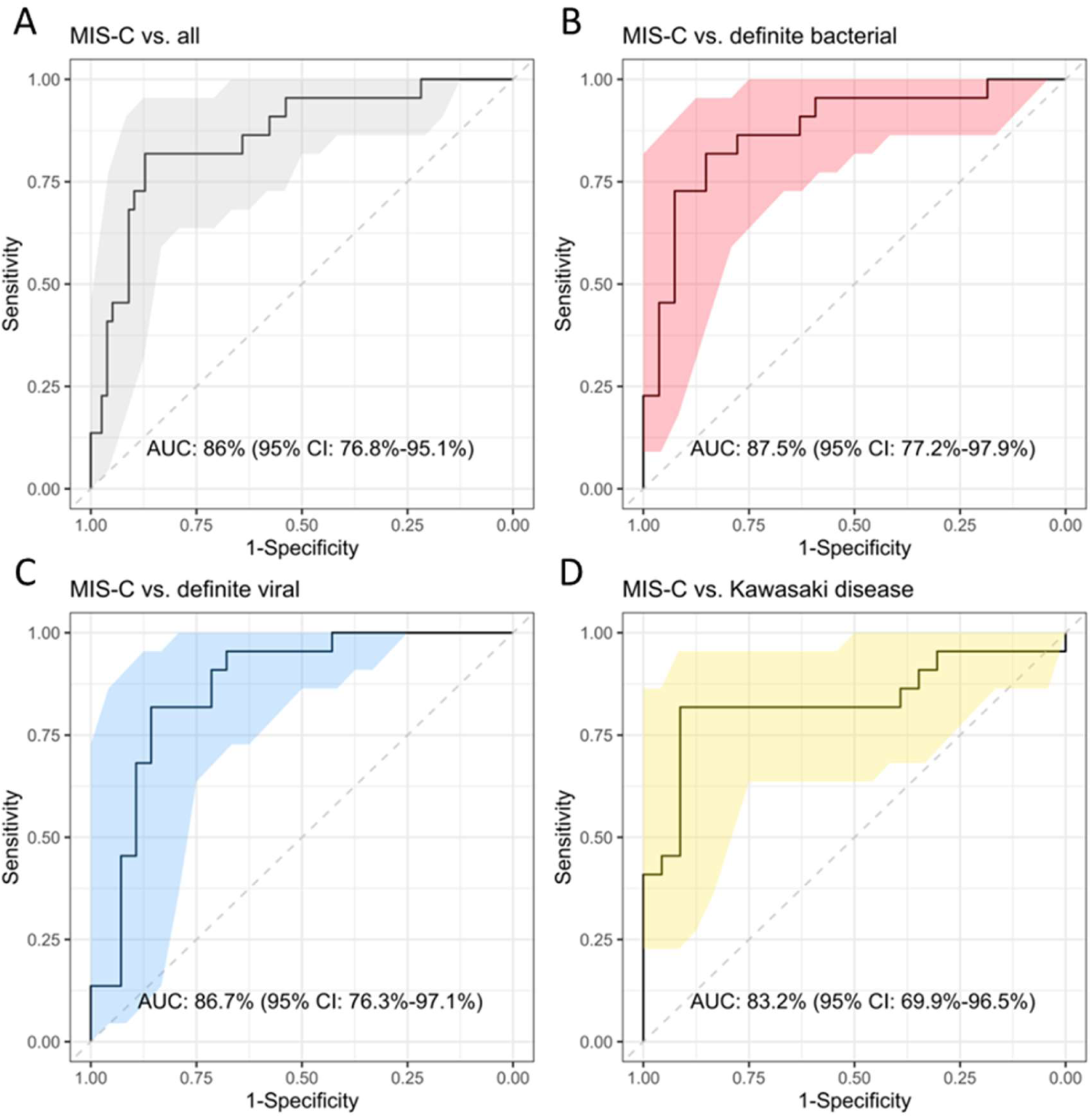
Receiver operating characteristic (ROC) curves visualising the performance of the 3-protein signature (PCSK9, CD163 and CXCL9). Area under the ROC curve (AUC) and 95% confidence intervals (CI) are shown on the plots.

When broken down into pairwise comparisons, the 3-protein signature showed the best performance for MIS-C *vs*. DB with an AUC of 87.5% (95% CI: 77.2%-97.9%; Figure 2B), followed by MIS-C *vs*. DV with an AUC of 86.7% (95% CI: 76.3%-97.1%; Figure 2C), and then MIS-C *vs*. KD with an AUC of 83.2% (95% CI: 69.9%-96.5%; Figure 2D).

The performance of smaller combinations of proteins was also evaluated to determine whether all three markers (PCSK9, CD163, and CXCL9) were required in the model (Table S3). The combination of all 3 proteins was optimal for distinguishing MIS-C from all groups combined (DB, DV, KD), and also for MIS-C *vs*. DB. For MIS-C *vs*. DV and MIS-C *vs*. KD, CD163+PCSK9 demonstrated the best performance. Despite this improved performance for these comparisons, the 3-protein signature was taken forward for subsequent analyses given the importance in distinguishing MIS-C from severe bacterial infections and the implications for treatment administration.

C-reactive protein (CRP) is a widely measured biomarker, often used for identifying bacterial infections (30). Given the widespread availability of CRP measurement, we evaluated whether adding CRP to the 3-protein model improved its diagnostic performance (Table S3). The 3-protein signature + CRP improved the performance for MIS-C *vs*. KD by 9%, however, a 16% reduction in performance was observed for MIS-C *vs*. DB. CRP could not be accurately assessed for the classification of MIS-C vs DV, as CRP was used in the initial clinical assignment of DV patients (DV patients were required to have CRP <60mg/L).

### Lower ARG1 and CORIN plasma levels are associated with severity in MIS-C

Associations between plasma protein concentrations and clinical variables were tested (Table 2). ARG1 was found to be associated with oxygen requirement (Table 2; Figure 3A; Bonferroni-adjusted p-value: 0.028) and inotrope requirement (Table 2; Figure 3B; Bonferroni-adjusted p-value: 0.020). Lower levels of ARG1 were observed in MIS-C patients requiring oxygen or inotrope administration, suggesting that reduced ARG1 concentration is indicative of elevated severity of MIS-C. CORIN concentration was associated with shock (Table 2; Figure 3C; Bonferroni-adjusted p-value: 0.029), following a similar trend with lower levels observed amongst patients who experienced shock. Where data was available, associations were also tested in DB, DV, and KD for ARG1 and inotrope requirement, oxygen requirement, and for CORIN and shock. No significant associations were observed.

**Figure 3.**
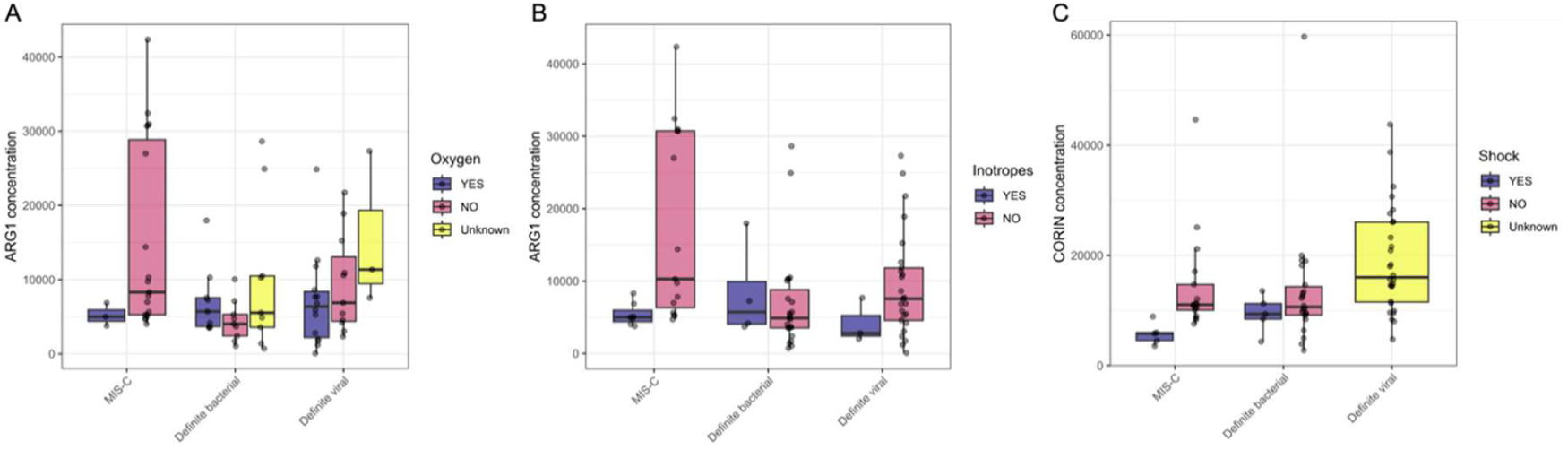
Levels of ARG1 and CORIN amongst patients stratified according to whether they required inotropes, required oxygen, or presented with shock.

**Table 2.** Bonferroni-adjusted p-values showing the association between clinical variables and each of the proteins measured in MIS-C patients. Associations between protein levels and PICU admission, oxygen requirement, non-invasive ventilation requirement, inotrope requirement, cardiac involvement, GI involvement, rash, and conjunctivitis were tested using two-sided t-tests, whilst the association between protein levels and BMI, days of symptoms, CRP, WBC, neutrophil, lymphocytes, monocytes, and platelet levels were tested using linear regression models. Bonferroni-adjusted p-values <0.05 are in bold, underlined font. P-values are adjusted for each clinical/laboratory variable.

| Proteins | PCSK9 | CD163 | CXCR9 | ARG1 | CORIN | MIP3A | ADAMTS2 |
| --- | --- | --- | --- | --- | --- | --- | --- |
| PICU admission | 1.000 | 1.000 | 1.000 | 0.077 | 0.537 | 1.000 | 1.000 |
| Oxygen requirement | 1.000 | 1.000 | 1.000 | **<u>0.028</u>** | 0.092 | 1.000 | 1.000 |
| Inotrope requirement | 0.103 | 0.593 | 0.652 | **<u>0.020</u>** | 0.372 | 1.000 | 1.000 |
| Non-invasive ventilation requirement | 1.000 | 1.000 | 0.560 | 0.063 | 0.820 | 1.000 | 1.000 |
| Cardiac involvement | 0.862 | 1.000 | 1.000 | 0.079 | 0.994 | 1.000 | 1.000 |
| GI involvement | 1.000 | 1.000 | 1.000 | 0.344 | 1.000 | 1.000 | 1.000 |
| Rash | 1.000 | 1.000 | 1.000 | 1.000 | 1.000 | 1.000 | 1.000 |
| Shock | 1.000 | 1.000 | 1.000 | 0.058 | **<u>0.029</u>** | 1.000 | 1.000 |
| Conjunctivitis | 0.837 | 1.000 | 1.000 | 1.000 | 0.716 | 0.565 | 1.000 |
| Days of symptoms | 1.000 | 1.000 | 1.000 | 1.000 | 1.000 | 1.000 | 1.000 |
| CRP levels | 0.678 | 0.437 | 0.437 | 0.678 | 0.437 | 0.229 | 0.698 |
| WBC levels | 1.000 | 1.000 | 0.942 | 1.000 | 1.000 | 1.000 | 1.000 |
| Neutrophil levels | 1.000 | 1.000 | 0.937 | 1.000 | 1.000 | 1.000 | 1.000 |
| Lymphocytes | 1.000 | 1.000 | 1.000 | 1.000 | 0.207 | 1.000 | 1.000 |
| Monocytes | 1.000 | 1.000 | 1.000 | 1.000 | 0.980 | 1.000 | 1.000 |
| Platelets | 1.000 | 1.000 | 1.000 | 1.000 | 0.157 | 1.000 | 1.000 |

## DISCUSSION

Weeks after SARS-CoV-2 infection, some children developed a hyperinflammatory condition termed Multisystem Inflammatory Syndrome in Children (MIS-C). The clinical presentation and laboratory findings in MIS-C are similar to other inflammatory and infectious diseases such as severe bacterial or viral infection and Kawasaki Disease, which makes the appropriate diagnosis and treatment of MIS-C challenging.

Here, we evaluated seven candidate host protein biomarkers in a cohort of 108 children with MIS-C, KD, definite bacterial infections (DB), definite viral infections (DV), and healthy controls (HC). All MIS-C cases met the WHO case definitions and over half were admitted to PICU. These patients were compared to KD, DB, and DV cases recruited prior to the COVID-19 pandemic. Five of the candidate biomarkers were selected from RNA-Seq and two from the literature because of their reported roles in the gastrointestinal involvement and hyper-inflammatory state observed early in the course of MIS-C disease, respectively (6,17,28).

The results showed that three proteins (PCSK9, CD163, CXCL9) were significantly higher in MIS-C cases when compared to the other comparator groups. More importantly, the performance of PCSK9, CD163, and CXCL9 when combined into a 3-protein signature could distinguish MIS-C patients from our other disease controls with an AUC of 86.0% (95% CI,76.8%-95.1%). When we included CRP levels into the 3-protein model, we observed an increase in performance for distinguishing MIS-C from KD of 9%. Given the overlap in clinical features between MIS-C and KD, especially in incomplete KD cases, these findings could be clinically useful due to the importance of timely administration of IVIG to patients with suspected KD to prevent coronary artery aneurysm formation (31). Similarly, we observed an increase in distinguishing MIS-C from DV infections when including CRP levels into the 3-protein model, however, this finding needs to be further validated in an independent cohort comprised of patients with viral infections that have been phenotyped independent of CRP, as CRP levels were used in the clinical phenotyping of the DV cases in this study (i.e., <60 mg/L).

The proteins included in the 3-protein signature may have biological functions that can provide insight into the pathogenesis of MIS-C. CD163 is a transmembrane macrophage-specific hemoglobin-haptoglobin scavenger receptor that is elevated in MAS and other vasculitic conditions. The biomarker form of this protein is the soluble CD163 (sCD163) found in plasma and produced from increased sCD163 shedding mediated by TNF-α (32). Increased abundance of sCD163 and marked elevation of macrophage activity have been recognized as markers in several inflammatory diseases (33). Mostafa and colleagues (34) reported increased levels of sCD163 in children with SAR-CoV-2 infection and MIS-C, compared to healthy controls, which likely reflects the exaggerated pro-inflammatory host response and suggests a potential therapeutic role for sCD163 antagonists. CD163 levels have also been shown to strongly correlate with the marked neutrophilia observed in MIS-C, suggesting this is a result of macrophage activation and supporting the observation that the highest levels of this protein are found in the most severe cases of MIS-C (20,35). CXCL9 is one of three chemokines that selectively bind to the C-X-C Motif Chemokine Receptor 3 (CXCR3) and its expression is primarily driven by IFN-γ (36). CXCL9 is commonly expressed by peripheral blood mononuclear cells, but more specifically by macrophages, and is best known for its role in the inflammatory response by mediating immune cell migration and activation (37). CXCL9 and its mediator, IFN-γ, have been reported to be more abundant in MIS-C cases than in children with mild and severe COVID, confirming the hyperinflammatory state observed in MIS-C (17,20,28). Moreover, our results support these findings and validate CD163 and CXCL9 as viable biomarkers for the diagnosis of MIS-C when compared to KD or other common bacterial and viral childhood infections.

Higher levels of PCSK9 were observed in MIS-C cases when compared to all other disease comparator groups. PCSK9 is vital in the metabolism of plasma cholesterol by regulating the levels of low-density lipoproteins (LDL) receptors which filter out the cholesterol-rich LDL particles from plasma (38,39,40). Increased PCSK9 expression has been linked to lower levels of LDL receptors and consequently higher LDL levels. Cholesterol is essential for SARS-CoV-2 infection as the virus binds to cell surface Angiotensin Converting Enzyme 2 (ACE2) fused to cholesterol, causing a signaling cascade allowing the lipid-enveloped RNA to enter host cells (41,42). In systemic inflammatory conditions, disruption of the vascular endothelium can lead to dysregulation of lipid transport and metabolism and an increase in circulating LDL and PCSK9. This explains why higher cholesterol levels are associated with higher susceptibility to SARS-CoV-2 (42). PCSK9 also exhibits a pro-inflammatory effect by promoting TNF-α expression while suppressing the anti-inflammatory markers ARG1 and IL-10 (43). The beneficial effect of using PCSK9 inhibitors in adults with severe COVID-19 has been demonstrated (44,45,46), however, elevated levels of this protein in MIS-C cases have, to our knowledge, not been previously reported.

ARG1 was not found to be significantly different between our MIS-C cases and other comparator groups, however, levels of this protein were significantly lower in the most severe MIS-C cases. Moreover, lower levels of ARG1 were associated with severe cases of MIS-C requiring oxygen and inotropes. ARG1 is the final enzyme involved in the urea cycle, hydrolyzing arginine to urea, and as such, is highly expressed in the liver (47,48). It also plays an important role in the immune response where it is released extracellularly under inflammatory conditions to inhibit inflammatory damage and immunity towards intracellular pathogens by reducing T-cell proliferation and cytokine production (49,50,51). ARG1 dysregulation has become synonymous with various pathological processes involving cardiovascular, immunological, neurodegenerative, and tumorigenic disorders (52). Elevated levels of ARG1 have been reported in COVID-19 cases with high viral loads (51,53) so it is possible that the high levels of PCSK9 observed in severe MIS-C cases are suppressing ARG1.

Lower levels of CORIN were also associated with severe cases with shock. CORIN converts pro-atrial natriuretic peptide (pro-ANP) to biologically active ANP, a cardiac hormone that regulates blood volume and pressure by reducing plasma volume by renal excretion of salt and water, vasodilation, and increased vascular permeability (54). Higher pro-ANP levels, which could result from lower levels of CORIN, are associated with poor survival prognosis in children and adults with severe sepsis and septic shock (55,56). The lower levels of CORIN we observed could therefore result in higher levels of pro-ANP and lower levels of ANP, which could contribute to the hypertension, inflammation, and cardiac manifestations associated with severity and shock in MIS-C.

We did not observe significant differences in the levels of ADAMTS2 or CCL20 in the MIS-C cases when compared to our other disease group. ADAMTS2, a biomarker we selected from RNA data, is a metalloprotease that processes procollagen to provide strength and support to many body tissues. Subsequent to our screening of RNA-Seq data, two other groups have reported increased gene expression of ADAMTS2 in COVID-19 and MIS-C cases, however, these groups were compared to healthy controls rather than other disease groups (57,58). Similarly, Gruber et al., (59) reported an increased abundance of CCL20 in patients with MIS-C. Our inability to replicate their results could be due to the difference in the control groups that were used: KD, DB, and DV as compared to non-ICU COVID-19-infected children, young adults, adults, and convalescent adults.

Although our study provides useful information on potential protein biomarkers for MIS-C, there are several limitations that should be taken into consideration. Firstly, our sample size for MIS-C (n=22) was restricted by the number of children that had been recruited at the time of this study that met all WHO criteria. Secondly, these patients were recruited during the period of April 2020-August 2021 and, due to the variants that were circulating during this period, would not include the Omicron variant that was first detected in November 2021. Further investigation of the biomarkers we present in this study is required in a larger independent cohort that includes other relevant DB and DV groups with overlapping clinical features of MIS-C including TSS, Macrophage Activation Syndrome (MAS), and Hemophagocytic Lymphohistiocytosis (HLH). Furthermore, our MIS-C cases were older than our other disease comparator groups and the performance of our signature should be tested in younger children with MIS-C. A limitation of all diagnostic studies comparing MIS-C and KD is the lack of a diagnostic test for either disease, however, we included KD patients that were recruited pre-pandemic to avoid errors in phenotyping. None of the KD patients in this study required inotropes and were, therefore, less severe than the MIS-C, DB, and DV groups. The severity associations that we observed for ARG1 and CORIN require further validation in more severe KD cases or KD shock to confirm if this finding is unique to MIS-C.

Finally, this study reports the first protein-based diagnostic signature to discriminate MIS-C from KD and other common bacterial and viral infections. Our results show that the increase in specific proteins involved in macrophage activation, endothelial dysfunction, and lipid metabolism can discriminate MIS-C from other inflammatory diseases in children. We further explored the associations between plasma levels and clinical outcomes to find both ARG1 and CORIN levels are lower in severe MIS-C. Once validated, our results could form the foundation for the development of a point-of-care diagnostic test that could assist pediatricians to diagnose and determine the best course of treatment for MIS-C.

## Data Availability

All data produced in the present study are available upon reasonable request to the authors.

## ACKNOWLEDGEMENTS

We are grateful to all patients and families that contributed towards this study. We would like to thank the UK KD Genetic consortium and the PERFORM and DIAMONDS consortia for their contributions to obtaining funding, patient recruitment, clinical data collection and entry, assignment of patient phenotypes, and processing and storage of research samples used in this analysis (see all Consortia members in Supplementary Text File 1). We would like to acknowledge our clinical collaborators across the UK who, with support from the UK NIHR Clinical Research Network, rapidly established recruitment of children under difficult circumstances during the COVID pandemic, including the MIS-C patients reported in this paper.

## AUTHOR CONTRIBUTIONS

**Conceptualization**: MK, JH, AC, ML, SH

**Methodology:** MK, DER, IK, SY, HJ, SH

**Validation:** SY, DER, IK, SH

**Formal Analysis:** SY, SH, HJ, MK

**Original draft preparation:** SY, MK, ML, DER, IK, HJ, JH, RG, SC, AC, SH

**Reviewing and Editing:** LC, PS, PA, RB, EC, ME, CF, TK, FMT, MMT, SP, MP, PR, LR, LS, NS, CFS, MT, EU, MVF, CV, UVB, SY, DZ

**Recruited and phenotyped patients used in this study:** PS, RG, SC, JH, AC, PA, RB, EC, ME, CF, TK, FMT, SP, MP, PR, LR, LS, NS, CFS, MT, EU, MVF, CV, UVB, SY, DZ

**Acquisition of funding:** HJ, RG, PS, PA, RB, EC, ME, CF, TK, FMT, MMT, SP, MP, PR, LR, LS, NS, CFS, MT, EU, MVF, CV, UVB, SY, DZ, LC, AC, JH, MK, SH, ML

## FUNDING

This work was supported by the European Union’s Horizon 2020 Program under grants (848196 DIAMONDS, 668303 PERFORM, [ML]) and the UK National Institute for Health Research Imperial Biomedical Research Centre at Imperial College (WMNP_P69099 [JH and ML]), the Javon Charitable Trust (ML) and the Children of St Mary’s Intensive Care Kawasaki Disease Research Fund (JH and ML).

## CONFLICT OF INTEREST

The authors declare no conflict of interest. The funders had no role in the design of the study; in the collection, analyses, or interpretation of data; in the writing of the manuscript, or in the decision to publish the results.

## Notes

### Competing Interest Statement

The authors have declared no competing interest.

### Funding Statement

This work was supported by the European Union’s Horizon 2020 Program under grants (848196 DIAMONDS, 668303 PERFORM) and the UK National Institute for Health Research Imperial Biomedical Research Centre at Imperial College (WMNP_P69099), the Javon Charitable Trust and the Children of St Mary's Intensive Care Kawasaki Disease Research Fund.

### Author Declarations

Informed consent was taken from all patients recruited to the study. Recruitment at all locations took place after ethical permissions were in place for that area. In the DIAMONDS study (Diagnosis and Management of Febrile Illness using RNA Personalized Molecular Signature Diagnosis https://www.diamonds2020.eu) and the PERFORM study (Personalized Risk assessment in Febrile illness to Optimize Real-life Management, perform2020.org/), the consortium agreed on a finalized protocol and supporting documents that were translated into local languages. Each participating country took responsibility for gaining ethical approval in their region. For DIAMONDS, the lead site received ethical approval for United Kingdom centers from the London-Dulwich research ethics committee (20/HRA/1714). For the PERFORM study, the lead site received approval ethical approval for United Kingdom centers from the London-Central research ethics committee (16/LO/1684). The Genetic Determinants of Kawasaki Disease for Susceptibility and Outcome study recruited in the United Kingdom only, and ethical approval was granted by the London-Fulham research ethics committee (13/LO/0026). During the conduct of the study, clinical data and samples were identified only by anonymized study numbers.

