## Supplemental Material for "Plasma protein biomarkers distinguish Multisystem Inflammatory Syndrome in Children (MIS-C) from other pediatric infectious and inflammatory diseases"

### Supplementary Figures

***Figure S1.*** *Breakdown of causative agents identified in the Definite Bacterial and Definite Viral groups.*

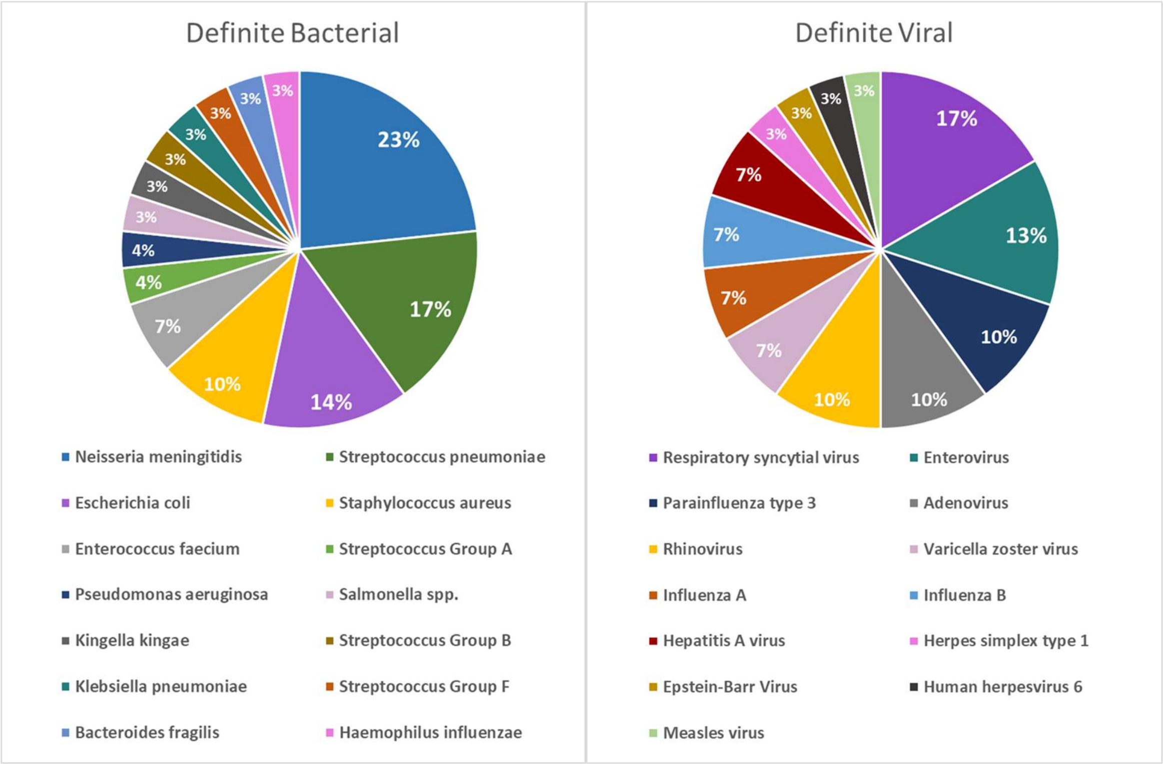

***Figure S2.*** *Principal component analysis (PCA) biplots generated from the protein abundance values of the 7 proteins measured in this study. Principal component (PC) 1 and 2 are shown. Points are coloured by disease group (A), sex (B), and age in years (C).*

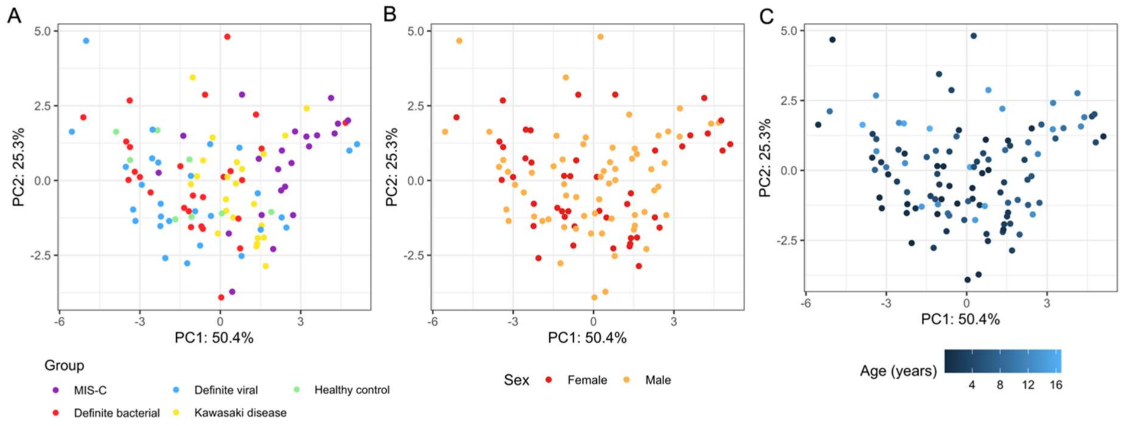

**Supplementary Tables**

***Table S1.*** *Proteins selected for analysis, their function and reason for inclusion. LFC= Log-fold change. P= adjusted p-value.*

| Protein | UniProt ID | Function | Justification for inclusion | Relevant publication(s) |
|---|---|---|---|---|
| ADAMTS2 | O95450 | Metalloprotease cleaving procollagens | RNA-Seq analysis: LFC 3.42, p= 3.03 E-09 | |
| ARG1 | O95450 | Hydrolase in urea cycle | RNA-Seq analysis: LFC 2.31, p= 9.47 E-12 | |
| CCL20 | P78556 | Inflammation and homing chemokine | Literature search | (6) |
| CD163 | Q86VB7 | Haemoglobin scavenger receptor | RNA-Seq analysis: LFC 2.66, p=4.34 E-18 | |
| CORIN | Q9Y5Q5 | Serine protease responsible atrial natriuretic peptide (ANP) production | RNA-Seq analysis: LFC 2.75, p= 3.79 E-20 | |
| CXCL9 | Q07325 | Chemokine involved in inflammatory response | Literature search | (28, 17) |
| PCSK9 | Q8NBP7 | Low-density lipoprotein (LDL) receptor regulation | RNA-Seq analysis: LFC 4.94, p= 3.10 E-08 | |

***Table S2.*** *Significance levels of each of the proteins measured. P-values obtained from the Tukey test for the pairwise comparisons and ANOVA for MIS-C vs. all groups, with p-values corrected using the Bonferroni adjustment. P-values from the Tukey test are corrected as part of the Tukey test. MIS-C = multisystem inflammatory syndrome in children; KD = Kawasaki disease; DB = definite bacterial; DV = definite viral; HC = healthy control. Significant p-values are shown in underlined and bold font.*

| Protein | MIS-C *vs*. KD, DB, DV, HC | MIS-C *vs*. KD | MIS-C *vs*. DB | MIS-C *vs*. DV |
|---|---|---|---|---|
| **PCSK9** | **$\underline{3.34 \times 10^{-5}}$** | **$\underline{1.25 \times 10^{-4}}$** | **<u>0.001</u>** | **$\underline{3.4 \times 10^{-4}}$** |
| **CD163** | **$\underline{1.43 \times 10^{-4}}$** | **<u>0.016</u>** | **<u>0.018</u>** | **$\underline{5.11 \times 10^{-5}}$** |
| **CXCL9** | **<u>0.008</u>** | **<u>0.017</u>** | **<u>0.004</u>** | 0.050 |
| **ARG1** | 0.122 | 0.918 | 0.081 | 0.311 |
| **CORIN** | 0.105 | 0.531 | 0.999 | 0.097 |
| **CCL20** | 0.385 | 0.934 | 0.691 | 0.999 |
| **ADAMTS2** | 0.105 | 0.999 | 0.926 | 0.451 |

***Table S3.*** *The performance of all possible combinations of PCSK9, CD163 and CXCL9 in distinguishing MIS-C from DB, DV and KD. *: “All” includes DB, DV and KD. AUCs and 95% confidence intervals are shown in the table. **: for this comparison, MIS-C vs. all represents MIS-C vs. DB and KD as CRP was used in the classification of DV patients. ***: perfect classification is expected as CRP was used in the classification of DV samples.*

| Combination | MIS-C *vs.* all* | MIS-C *vs.* DB | MIS-C *vs.* DV | MIS-C *vs.* KD |
|---|---|---|---|---|
| PCSK9+CD163+ CXCL9 | 86.9% (76.8%- 95.1%) | 87.5% (77.2%- 97.9% | 86.7% (76.3%- 97.1%) | 83.2% (69.9%- 96.5) |
| CD163+PCSK9 | 85.0% (75.7%- 94.3%) | 81.8% (69.1%- 94.5%) | 88.8% (79.7%- 97.9%) | 84.0% (72.1%- 95.9%) |
| CD163+CXCL9 | 84.3% (75.0%- 93.7%) | 86.5% (75.9%- 97.2%) | 86.4% (75.9%- 96.9%) | 79.3% (64.2%- 94.3%) |
| PCSK9+CXCL9 | 82.6% (72.0%- 93.2%) | 86.5% (76.2%- 96.9%) | 82.5% (70.4%- 94.5%) | 78.1% (62.9%- 93.2%) |
| PCSK9 | 74.2% (61.3%- 87.2%) | 69.7% (54.4%- 85.0%) | 75.3% (61.1%- 89.5%) | 78.3% (64.4%- 92.1%) |
| CD163 | 82.8% (74.0%- 91.5%) | 81.1% (68.5%- 93.8%) | 88.8% (79.8%- 97.8%) | 77.3% (63.5%- 91.0%) |
| CXCL9 | 81.4% (71.0%- 91.9%) | 85.0% (74.1%- 96.0%) | 82.8% (71.0%- 94.6%) | 75.5% (59.5%- 91.5%) |
| 3-proteins + CRP** | 81.1% (71.3%- 90.9%) | 71.5% (56.9%- 86.2%) | 100% *** | 92.3% (84.8%- 99.8%) |

**Supplementary Text File 1**

**DIAMONDS, PERFORM and UK Kawasaki Disease Genetic Consortia**

### 1.1. DIAMONDS Consortium

https://www.diamonds2020.eu/

**PARTNER: Imperial College (Coordinating Centre) (UK)**

*Chief investigator/DIAMONDS coordinator:*

Michael Levin[1]

*Principal and co-investigators (alphabetical order)[1]*

Aubrey Cunnington; Jethro Herberg; Myrsini Kaforou; Victoria Wright

*Section of Paediatric Infectious Diseases Research Group (alphabetical order)[1]*

Evangelos Bellos; Claire Broderick; Samuel Channon-Wells; Samantha Cooray; Tisham De (database work package lead); Giselle D'Souza; Leire Estramiana Elorrieta; Diego Estrada-Rivadeneyra; Rachel Galassini (Clinical Trial Manager); Dominic Habgood-Coote; Shea Hamilton (Proteomics); Heather Jackson; James Kavanagh; Mahdi Moradi Marjaneh; Stephanie Menikou; Samuel Nichols; Ruud Nijman; Harsita Patel; Ivana Pennisi; Oliver Powell; Ruth Reid; Priyen Shah; Ortensia Vito; Elizabeth Whittaker; Clare Wilson; Rebecca Womersley

*Recruitment team at Imperial College Healthcare NHS Trust, London (alphabetical order)[2]*

Amina Abdulla; Sarah Darnell; Sobia Mustafa

*Engineering Team*

Pantelis Georgiou[3] (engineering lead); Jesus-Rodriguez Manzano[4]; Nicolas Moser[3]; Ivana Pennisi[1]

[1]Section of Paediatric Infectious Disease, Imperial College London, Norfolk Place, London W2 1PG, UK

[2]Children's Clinical Research Unit, St Mary's Hospital, Praed Street, London W2 1NY, UK

[3] Imperial College London, Department of Electrical and Electronic Engineering, South Kensington Campus, London, SW7 2AZ, UK

[4] Imperial College London, Department of Infectious Disease, Section of Adult Infectious Disease, Hammersmith Campus, London, W12 0NN, UK

**UK Non-Consortium Clinical Recruiting Sites**

*Evelina London Children's Hospital, Guy's and St Thomas' NHS Foundation Trust; King's College London [combined]*

Michael Carter[1,2] (principal investigator); Shane Tibby[1,2] (co-investigator)

*Recruitment team (alphabetical order):* Jonathan Cohen[1]; Francesca Davis[1;] Julia Kenny[1]; Paul Wellman[1]; Marie White[1]

*Laboratory team (alphabetical order):* Matthew Fish[3]; Aislinn Jennings[4]; Manu Shankar-Hari[3,4]

[1] Evelina London Children's Hospital, Guy's and St Thomas' NHS Foundation Trust, London, UK

[2] Department of Women and Children's Health, School of Life Course Sciences, King's College London, UK

[3] Department of Infectious Diseases, School of Immunology and Microbial Sciences, King's College London, London, UK

[4] Department of Intensive Care Medicine, Guy's and St Thomas' NHS Foundation Trust, London, UK

*University Hospitals Sussex*

Katy Fidler[1] (principal investigator); Dan Agranoff[2] (co-investigator)

*Recruitment team*; Vivien Richmond[1,3], Mathhew Seal[2]

[1] Royal Alexandra Children's Hospital, University Hospitals Sussex, Brighton, UK

[2] Dept of Infectious Diseases, University Hospitals Sussex, Brighton, UK

[3] Research Nurse team, University Hospitals Sussex, Brighton, UK

<u>*University Hospital Southampton NHS Foundation Trust*</u>

Saul Faust[1] (principal investigator); Dan Owen[1] (co-investigator);

*Recruitment team*; Ruth Ensom[2]; Sarah McKay[2]; Diana Mondo[3], Mariya Shaji[3]; Rachel Schranz[3] *(alphabetical order)*

[1] NIHR Southampton Clinical Research Facility, University Hospital Southampton NHS Foundation Trust and University of Southampton, UK

[2] NIHR Southampton Clinical Research Facility, University Hospital Southampton NHS Foundation Trust, UK

[3] Department of R&D, University Hospital Southampton NHS Foundation Trust, UK

<u>*Barts Health NHS Trust*</u>

Prita Rughnani[1, 2, 3] (principal investigator 2020-2021); Amutha Anpananthar[1, 2, 3] (principal investigator 2021-to date); Susan Liebeschuetz[2] (co-investigator), Anna Riddell[1] (co-investigator)

*Recruitment team;* Nosheen Khalid[1, 3,]Ivone Lancoma Malcolm, Teresa Simagan[3] *(alphabetical order)*

[1] Royal London Hospital, Whitechapel Rd, London E1 1FR, UK

[2] Newham University Hospital, Glen Rd, London E13 8SL, UK

[3] Whipps Cross University Hospital, *Whipps Cross* Road, London, E11 1NR, UK

<u>*Great Ormond Street Hospital for Children NHS Foundation Trust*</u>

Mark Peters[1,2] (principal investigator); Alasdair Bamford[1,2] (co-investigator)

*Recruitment team;* Lauran O'Neill[1]

[1] Great Ormond Street Hospital, London, WC1N 3JH, UK

[2] UCL Great Ormond St Institute of Child Health, WC1N 1EH, UK

*Cambridge University Hospitals NHS Foundation Trust*

Nazima Pathan[1,2] (principal investigator)

*Recruitment team; Esther Daubney[1], Deborah White[1] (alphabetical order)*

[1]Addenbrooke's Hospital, Hills Road, Cambridge CB2 0QQ, UK

[2]Department of Paediatrics, University of Cambridge, Cambridge CB2 0QQ, UK

*University College London Hospitals NHS Foundation Trust*

Melissa Heightman[1] (principal investigator); Sarah Eisen[1] (co-investigator)

*Recruitment team*; Terry Segal[1], Lucy Wellings[1] *(alphabetical order)*

[1] University College London Hospital, Euston Road, London NW1 2BU, UK

*St George's University Hospitals NHS Foundation Trust*

Simon B Drysdale[1] (principal investigator)

*Recruitment team; Nicole Branch[1], Lisa Hamzah[1], Heather Jarman[1] (alphabetical order)*

[1] St George's Hospital, Blackshaw Road, London SW17 0QT, UK

*Lewisham and Greenwich NHS Trust*

Maggie Nyirenda[1, 2,](principal investigator)

*Recruitment team* Lisa Capozzi[1], Emma Gardiner[1] *(alphabetical order)*

[1]University Hospital Lewisham, London SE13 6LH, UK

[2] Queen Elizabeth Hospital Greenwich, London SE18 4QH, UK

*Liverpool University Hospitals NHS Foundation Trust*

Robert Moots[1] (principal investigator); Magda Nasher[2] (principal investigator)

*Recruitment team*; Anita Hanson[2]; Michelle Linforth[1]

[1] Aintree University Hospital, Lower Lane, Liverpool L9 7AL, UK

[2] Royal Liverpool Hospital, Prescot St, Liverpool L7 8XP, UK

*Leeds Teaching Hospitals NHS Trust*

Sean O'Riordan[1] (principal investigator)

*Recruitment team*; Donna Ellis[1]

[1]Leeds Children's Hospital, Leeds LS1 3EX, UK

*King's College Hospital NHS Foundation Trust*

Akash Deep[1] (principal investigator)

*Recruitment team;* Ivan Caro[1]

[1] Kings College Hospital, Denmark Hill, London SE5 9RS, UK

*Sheffield Children's NHS Foundation Trust*

Fiona Shackley [1] (principal investigator);

*Recruitment team*; Arianna Bellini,[1] Stuart Gormley[1] *(alphabetical order)*

[1]Sheffield Children's Hospital, Broomhall, Sheffield S10 2TH, UK

*University Hospitals of Leicester NHS Foundation Trust*

Samira Neshat[1] (principal investigator)

[1]Leicester General Hospital, Leicester LE1 5WW, UK

*Birmingham Women's and Children's Hospital NHS Foundation Trust*

Barnaby J Scholefield[1] (principal investigator)

*Recruitment team; Ceri Robbins[1], Helen Winmill[1] (alphabetical order)*

[1] Birmingham Children's Hospital, Steelhouse Lane, Birmingham B4 6NH, UK

**PARTNER: University of Oxford (UK)**

**Children's Hospital, John Radcliffe Hospital, Oxford**

Principal Investigator

Stéphane C. Paulus[1,2,3]

Co-Principal Investigator

Andrew J. Pollard[1,2,3,4]

Co-investigators

Mark Anthony[1] (neonates)

Recruitment team

Sarah Hopton[1], Danielle Miller[1], Zoe Oliver[1], Sally Beer[1], Bryony Ward[1]

[1]John Radcliffe Hospital, Oxford University Hospitals NHS Foundation Trust, Oxford, UK

[2]Department of Paediatrics, University of Oxford, UK

[3]Oxford Vaccine Group, University of Oxford, UK

[4]NIHR Oxford Biomedical Research Centre, Oxford, UK

**University of Oxford, Nepal Site**

Principal Investigator

Shrijana Shrestha[1]

Co-Principal Investigator

Andrew J Pollard[2,3]

**Nepal Research Team**

Meeru Gurung[1]

Puja Amatya[1]

Bhishma Pokhrel[1]

Sanjeev Man Bijukchhe[1]

**Oxford Research Team**

Tim Lubinda[2]

Sarah Kelly[2]

Peter O'Reilly[2]

[1]Paediatric Research Unit, Patan Academy of Health Sciences, Kathmandu, Nepal.

[2]Oxford Vaccine Group, Department of Paediatrics, University of Oxford, Oxford, United Kingdom.

[3]NIHR Oxford Biomedical Research Centre, Oxford, United Kingdom.

**PARTNER: SERGAS (Spain)**

Principal Investigators

Federico Martinón-Torres[1]

Antonio Salas[1,2]

GENVIP RESEARCH GROUP (in alphabetical order):

Fernando Álvez González[1], Xabier Bello[1,2], Mirian Ben García[1], Sandra Carnota[1], Miriam Cebey-López[1], María José Curras-Tuala[1,2], Carlos Durán Suárez[1], Luisa García Vicente[1], Alberto Gómez-Carballa[1,2], Jose Gómez Rial[1], Pilar Leboráns Iglesias[1], Federico Martinón-Torres[1], Nazareth Martinón-Torres[1], José María Martinón Sánchez[1], Belén Mosquera Pérez[1], Jacobo Pardo-Seco[1,2], Lidia Piñeiro Rodríguez[1], Sara Pischedda[1,2], Sara Rey Vázquez[1], Irene Rivero Calle[1], Carmen Rodríguez-Tenreiro[1], Lorenzo Redondo-Collazo[1], Miguel Sadiki Ora[1], Antonio Salas[1,2], Sonia Serén Fernández[1], Cristina Serén Trasorras[1], Marisol Vilas Iglesias[1].

[1] Translational Pediatrics and Infectious Diseases, Pediatrics Department, Hospital Clínico Universitario de Santiago, Santiago de Compostela, Spain, and GENVIP Research Group

(www.genvip.org), Instituto de Investigación Sanitaria de Santiago, Universidad de Santiago de Compostela, Galicia, Spain.

[2] Unidade de Xenética, Departamento de Anatomía Patolóxica e Ciencias Forenses, Instituto de Ciencias Forenses, Facultade de Medicina, Universidade de Santiago de Compostela, and GenPop Research Group, Instituto de Investigaciones Sanitarias (IDIS), Hospital Clínico Universitario de Santiago, Galicia, Spain

[3] Fundación Pública Galega de Medicina Xenómica, Servizo Galego de Saúde (SERGAS), Instituto de Investigaciones Sanitarias (IDIS), and Grupo de Medicina Xenómica, Centro de Investigación Biomédica en Red de Enfermedades Raras (CIBERER), Universidade de Santiago de Compostela (USC), Santiago de Compostela, Spain

**PARTNER: Liverpool (UK)**

Principal Investigators

Enitan D Carrol[1,2,]

Research Group (in alphabetical order):

Elizabeth Cocklin[1], Aakash Khanijau[1], Rebecca Lenihan[1], Nadia Lewis-Burke[1]

Karen Newall[4], Sam Romaine[1], [1] Department of Clinical Infection, Microbiology and Immunology, University of Liverpool Institute of Infection, Veterinary and Ecological Sciences , Liverpool, England

[2] Alder Hey Children's Hospital, Department of Infectious Diseases, Eaton Road, Liverpool, L12 2AP

[3] [4]Alder Hey Children's Hospital, Clinical Research Business Unit, Eaton Road, Liverpool, L12 2AP

**PARTNER: NATIONAL AND KAPODISTRIAN UNIVERSITY OF ATHENS (Greece)**

Principal Investigator: Maria Tsolia[1]

Co-Investigator: Irini Eleftheriou[1]

PID Unit: Nikos Spyridis[1], Maria Tambouratzi[1]

Pediatric Rheumatology Unit: Despoina Maritsi[1]

Lab: Antonios Marmarinos[1], Marietta Xagorari[1]

*Recruitment teams:*

Adult COVID19- Infectious Diseases: Lourida Panagiota, Pefanis Aggelos[2]

Adult COVID19: Akinosoglou Karolina, Gogos Charalambos, Maragos Markos[3]

Adult Inflammatory Diseases-Oncology: Voulgarelis Michalis , Stergiou Ioanna[4]

[1]2nd Department of Pediatrics, National and Kapodistrian University of Athens (NKUA), Children's Hospital "P, and A. Kyriakou", Athens, Greece

[2]1st Department of Infectious Diseases, General Hospital "Sotiria"

[3]Pathology Department, University of Patras, General Hospital "Panagia i Voithia"

[4]Pathophysiology Department, Medical Faculty, National and Kapodistrian University of Athens (NKUA), General Hospital "Laiko"

**Newcastle upon Tyne Hospitals NHS Foundation Trust and Newcastle University (UK) combined**

Principal Investigator:

Marieke Emonts [1,2,3] (all activities)

Co-investigators

Emma Lim[2,3,6] (all activities)

John Isaacs[1] (adult inflammatory)

Recruitment team (alphabetical), datamanagers, and GNCH Research unit:

Kathryn Bell[4], Stephen Crulley[4], Daniel Fabian[4], Evelyn Thomson[4], Diane Wallia[4], Caroline Miller[4] , Ashley Bell[4]

PhD Students/medical staff DIAMONDS

Fabian J.S. van der Velden[1,2] (all activities), Geoff Shenton[7] (oncology), Ashley Price[8,9] (Adult COVID)

Students

Owen Treloar [1,2] (quality control, data management and analysis)

Daisy Thomas[1,2] (recruitment)

Author Affiliations:

[1] Translational and Clinical Research Institute, Newcastle University, Newcastle upon Tyne UK

[2]Great North Children's Hospital, Paediatric Immunology, Infectious Diseases & Allergy, Newcastle upon Tyne Hospitals NHS Foundation Trust, Newcastle upon Tyne, United Kingdom.

[3]NIHR Newcastle Biomedical Research Centre based at Newcastle upon Tyne Hospitals NHS Trust and Newcastle University, Westgate Rd, Newcastle upon Tyne NE4 5PL, United Kingdom

[4]Great North Children's Hospital, Research Unit, Newcastle upon Tyne Hospitals NHS Foundation Trust, Newcastle upon Tyne, United Kingdom.

[6]Population Health Sciences Institute, Newcastle University, Newcastle upon Tyne, UK

[7]Great North Children's Hospital, Paediatric Oncology, Newcastle upon Tyne Hospitals NHS Foundation Trust, Newcastle upon Tyne, United Kingdom.

[8]Department of Infection & Tropical Medicine, Newcastle upon Tyne Hospitals NHS Foundation Trust, Newcastle upon Tyne, United Kingdom

[9]NIHR Newcastle In Vitro Diagnostics Co-operative (Newcastle MIC), Newcastle upon Tyne, United Kingdom.

**Servicio Madrileño de Salud (SERMAS) - Fundación Biomédica del Hospital Universitario 12 de Octubre (FIB-H12O) (Spain)**

Principal Investigators

Pablo Rojo[1 3]

Cristina Epalza [1,2]

SERMAS/FIB-H120 team:

Serena Villaverde [1,], Sonia Márquez[2], Manuel Gijón [2], Fátima Machín[2], Laura Cabello[2], Irene Hernández[2], Lourdes Gutiérrez[2], Ángela Manzanares [1]

Author Affiliations:

[1] Servicio Madrileño de Salud (SERMAS),Pediatric Infectious Diseases Unit, Department of Pediatrics, Hospital Universitario 12 de Octubre, Madrid, Spain

[2]Fundación Biomédica del Hospital Universitario 12 de Octubre (FIB-H12O), Unidad Pediátrica de Investigación y Ensayos Clínicos (UPIC), Hospital Universitario 12 de Octubre, Instituto de Investigación Sanitaria Hospital 12 de Octubre (i+12), Madrid, Spain.

[3] Universidad Complutense de Madrid, Faculty of Medicine, Department of Pediatrics, Madrid, Spain.

**Amsterdam University Medical Center (Amsterdam UMC), University of Amsterdam**

Principal Investigator:

T.W. (Taco) Kuijpers MD PhD [1,2] (all activities)

Co-investigators

M. (Martijn) van de Kuip MD PhD [1] (infectious disease)

A.M. (Marceline) van Furth MD PhD [1] (infectious disease)

J.M. (Merlijn) van den Berg MD PhD [1] (inflammatory disease)

Hospital Team (all activities):

Giske Biesbroek MD PhD [1], Floris Verkuil MD (PhD student) [1], Carlijn (C.W.) van der Zee MD (start 1/8/2022, PhD student) [1]

Recruitment:

Dasja Pajkrt MD PhD [1], Michael Boele van Hensbroek MD PhD [1], Dieneke Schonenberg MD [1], Mariken Gruppen MD [1], Sietse Nagelkerke MD PhD [1,2], medical students

Laboratory Team:

Machiel H Jansen [1], Ines Goetschalckx (PhD student) [2]

Author Affiliations:

[1] Amsterdam UMC, Emma Children's Hospital, Dept of Pediatric Immunology, Rheumatology and Infectious Disease, University of Amsterdam, The Netherlands

[2] Sanquin, Dept of Molecular Hematology, University Medical Center, Amsterdam, The Netherlands

**Bambino Gesù Children's Hospital (Rome-Italy)**

Principal Investigator

Lorenza Romani 1

Maia De Luca 1

Recruitment Team

Sara Chiurchiù 1

Martina Di Giuseppe 1

Affiliation

1 Infectious Disease Unit, Academic Department of Pediatrics, Bambino Gesù Children's Hospital, IRCCS, Rome 00165, Italy

**ERASMUS MC-Sophia Children's Hospital**

*Principal Investigator*

Clementien L. Vermont[2]

*Research group*

Henriëtte A. Moll[1], Dorine M. Borensztajn[1], Nienke N. Hagedoorn, Chantal Tan [1], Joany Zachariasse [1], Medical students [1]

Additional investigator

W Dik [3]

[1] Erasmus MC-Sophia Children's Hospital, Department of General Paediatrics, Rotterdam, the Netherlands

[2] Erasmus MC-Sophia Children's Hospital, Department of Paediatric Infectious Diseases & Immunology, Rotterdam, the Netherlands

[3] Erasmus MC, Department of immunology, Rotterdam, the Netherlands

**TAIWAN**

Ching-Fen (Kitty), Shen

Division of Infectious Disease, Department of Pediatrics, National Cheng Kung University Tainan, Taiwan

**Riga Stradins University (Riga, Latvia)**

Principal Investigator:

Dace Zavadska [1,2] (all activities)

Co-investigators

Sniedze Laivacuma [1,3] (adult cohorts)

Recruitment team:

Aleksandra Rudzate [1,2], Diana Stoldere [1,2], Arta Barzdina [1,2], Elza Barzdina [1,2], Sniedze Laivacuma[1,3], Monta Madelane [1,3]

Laboratory

Dagne Gravele [2], Dace Svile[2]

Author Affiliations:

[1] Riga Stradins University, Riga, Latvia

[2] Children clinical university hospital, Riga, Latvia

[3] Riga East clinical university hospital, Riga, Latvia

**Assistance Publique - Hôpitaux de Paris**

Principal Investigator:

Romain Basmaci [1,2]

Co-investigator:

Noémie Lachaume [1]

Recruitment team:

Pauline Bories [1], Raja Ben Tkhayat [1], Laura Chériaux [1], Juraté Davoust [1], Kim-Thanh Ong [1], Marie Cotillon [1], Thibault de Groc [1], Sébastien Le [1], Nathalie Vergnault [1], Hélène Sée [1], Laure Cohen [1], Alice de Tugny [1], Nevena Danekova [1]

Author Affiliations:

[1] Service de Pédiatrie-Urgences, AP-HP, Hôpital Louis-Mourier, F-92700 Colombes, France

[2] Université Paris Cité, Inserm, IAME, F-75018 Paris, France

**BioMérieux**

Principal Investigator:

Marine Mommert-Tripon

Co-investigator:

Karen Brengel-Pesce

Author Affiliations:

bioMérieux - Open Innovation & Partnerships Department, Lyon, France

**University Medical Centre Ljubljana, Slovenia**

Principal Investigator: Marko Pokorn [1,2,3]

Co-Investigator: Mojca Kolnik[2]

Research Group (in alphabetical order):

Tadej Avčin[2,3], Tanja Avramoska[2], Natalija Bahovec[1], Petra Bogovič[1], Lidija Kitanovski[2,3], Mirijam Nahtigal[1], Lea Papst[1], Tina Plankar Srovin[1], Franc Strle[1,2], Anja Srpčič[2], Katarina Vincek[1].

Affiliations:

1. Department of Infectious diseases, University Medical Centre Ljubljana, Slovenia
2. University Children's Hospital, University Medical Centre Ljubljana, Slovenia
3. Faculty of Medicine, University of Ljubljana, Slovenia
4. Centre for Clinical research, University Medical Centre Ljubljana

**University Medical Center Utrecht, Utrecht, The Netherlands**

*Principal Investigator*

Michiel van der Flier[1,5] (Pediatric Infectious Diseases and Immunology)

*Co-investigators*

Wim J.E. Tissing[5] (Pediatric Oncology)

Roelie M. Wösten-van Asperen[2] (Pediatric Intensive Care Unit)

Sebastiaan J Vastert[3] (Pediatric Rheumatology)

Daniel C Vijlbrief[4] (Pediatric Neonatal Intensive Care)

Louis J. Bont[1,5] (Pediatric Infectious Diseases and Immunology)

Tom F.W. Wolfs [1,5] (Pediatric Infectious Diseases and Immunology)

*PhD student*

Coco R. Beudeker[1,5] (Pediatric Infectious Diseases and Immunology)

*Affiliations:*

1.Pediatric Infectious Diseases and Immunology, 2. Pediatric Intensive Care Unit 3. Pediatric Rheumatology 4. Pediatric Neonatal Intensive Care, Wilhelmina Children's Hospital, University Medical Center Utrecht, Utrecht, The Netherlands

5. Princess Maxima Center for Pediatric Oncology, Utrecht, The Netherlands

**PARTNER**: **University of Bern, Inselspital, Bern University Hospital, University of Bern (Switzerland)**

Principal Investigators (alphabetical)

Philipp Agyeman[1]

Luregn Schlapbach[2,3]

Co-Investigator

Christoph Aebi[1]

Recruitment team

Mariama Usman[1], Stefanie Schlüchter[1], Verena Wyss[1], Nina Schöbi[1], Elisa Zimmermann[2] PhD, Marion Meier[2], Kathrin Weber[2]

[1] Department of Pediatrics, Inselspital, Bern University Hospital, University of Bern, Switzerland

[2] Department of Intensive Care and Neonatology, and Children`s Research Center, University Children`s Hospital Zurich, Zurich, Switzerland

[3] Child Health Research Centre, The University of Queensland, Brisbane, Australia

**Swiss Pediatric Sepsis Study group**

Philipp Agyeman, MD [1], Luregn J Schlapbach, MD, FCICM [2,3], Eric Giannoni, MD [4,5], Martin Stocker, MD [6], Klara M Posfay-Barbe, MD [7], Ulrich Heininger, MD [8], Sara Bernhard-Stirnemann, MD [9], Anita Niederer-Loher, MD [10], Christian Kahlert, MD [10], Giancarlo Natalucci, MD [11], Christa Relly, MD [12], Thomas Riedel, MD [13], Christoph Aebi, MD [1], Christoph Berger, MD [12]

**Affiliations:**

[1] Department of Pediatrics, Inselspital, Bern University Hospital, University of Bern, Switzerland

[2] Department of Intensive Care and Neonatology, and Children`s Research Center, University Children`s Hospital Zurich, Zurich, Switzerland

[3] Child Health Research Centre, The University of Queensland, Brisbane, Australia

[4] Clinic of Neonatology, Department Mother-Woman-Child, Lausanne University Hospital and University of Lausanne, Switzerland

[5] Infectious Diseases Service, Department of Medicine, Lausanne University Hospital and University of Lausanne, Switzerland

[6] Department of Pediatrics, Children’s Hospital Lucerne, Lucerne, Switzerland

[7] Pediatric Infectious Diseases Unit, Children’s Hospital of Geneva, University Hospitals of Geneva, Geneva, Switzerland

[8] Infectious Diseases and Vaccinology, University of Basel Children’s Hospital, Basel, Switzerland

[9] Children’s Hospital Aarau, Aarau, Switzerland

[10] Division of Infectious Diseases and Hospital Epidemiology, Children’s Hospital of Eastern Switzerland St. Gallen, St. Gallen, Switzerland

[11] Department of Neonatology, University Hospital Zurich, Zurich, Switzerland

[12] Division of Infectious Diseases and Hospital Epidemiology, and Children's Research Center, University Children's Hospital Zurich, Switzerland

[13] Children's Hospital Chur, Chur, Switzerland

**Micropathology Ltd (UK)**

Micropathology Ltd, The Venture Center, University of Warwick Science Park, Sir William Lyons Road, Coventry, CV4 7EZ

Principle Investigator; Prof Colin Fink

Co Investigators: Marie Voice, Leo Calvo-Bado, Michael Steele, Jennifer Holden

Research group: Benjamin Evans, Jake Stevens, Peter Matthews, Kyle Billing

**Medical University of Graz, Austria (MUG)**

Principal Investigator:

Werner Zenz[1] (all activities)

Co-investigators (in alphabetical order):

Alexander Binder[1] (grant application)

Benno Kohlmaier[1] (study design, recruitment)

Daniela S. Kohlfürst[1] (study design)

Nina A. Schweintzger[1] (all activities)

Christoph Zurl[1] (study design, recruitment)

Recruitment team, data managers, laboratory work (in alphabetical order):

Susanne Hösele[1], Manuel Leitner[1], Lena Pölz[1], Alexandra Rusu[1], Glorija Rajic[1], Bianca Stoiser[1], Martina Strempfl[1,] Manfred G. Sagmeister[1]

Clinical recruitment partners (in alphabetical order):

Sebastian Bauchinger[1], Martin Benesch[3], Astrid Ceolotto[1], Ernst Eber[2], Siegfried Gallistl[1], Harald Haidl[1], Almuthe Hauer[1], Christa Hude[1], Andreas Kapper[7], Markus Keldorfer[5], Sabine

Löffler[5], Tobias Niedrist[6], Heidemarie Pilch[5], Andreas Pfleger[2], Klaus Pfurtscheller[4], Siegfried Rödl[4], Andrea Skrabl-Baumgartner[1], Volker Strenger[3], Elmar Wallner[7]

Author Affiliations:

[1] Department of Pediatrics and Adolescent Medicine, Division of General Pediatrics, Medical University of Graz, Graz, Austria

[2]Department of Pediatric Pulmonology, Medical University of Graz, Graz, Austria

[3]Department of Pediatric Hematooncology, Medical University of Graz, Graz, Austria

[4]Paediatric Intensive Care Unit, Medical University of Graz, Graz, Austria

[5]University Clinic of Pediatrics and Adolescent Medicine Graz, Medical University Graz, Graz, Austria

[6]Clinical Institute of Medical and Chemical Laboratory Diagnostics, Medical University Graz, Graz, Austria

[7]Department of Internal Medicine, State Hospital Graz II, Location West, Graz, Austria

**SkylineDX**

Principle investigator: Dennie Tempel [1]

Co-investigators: Danielle van Keulen[1], Annelieke M Strijbosch [1],

Author affiliations:

[1] SkylineDx, Rotterdam, The Netherlands

**Project partner BBMRI-ERIC**

Maike K. Tauchert

Author affiliation:

Biobanking and BioMolecular Resources Research Infrastructure - European Research Infrastructure Consortium (BBMRI-ERIC), Neue Stiftingtalstrasse 2/B/6, 8010, Graz, Austria

**LMU Munich Partner (Germany)**

Principal Investigator:

Ulrich von Both[1,2] MD, FRCPCH (all activities)

Research group:

Laura Kolberg[1] MSc (all activities)

Patricia Schmied[1] (Study physician), Irene Alba-Alejandre[3] MD (Study physician)

Clinical recruitment partners (in alphabetical order):

Katharina Danhauser, MD[6], Nikolaus Haas, MD[11], Florian Hoffmann, MD[10], Matthias Griese, MD[7], Tobias Feuchtinger, MD[5], Sabrina Juranek, MD[4], Matthias Kappler, MD[7], Eberhard Lurz, MD[8], Esther Maier, MD[4], Karl Reiter, MD[10], Carola Schoen, MD[10], Sebastian Schroepf, MD[9]

Author Affiliations:

[1] Division of Pediatric Infectious Diseases, Department of Pediatrics, Dr. von Hauner Children's Hospital, University Hospital, LMU Munich, Munich, Germany

[2] German Center for Infection Research (DZIF), Partner Site Munich, Munich, Germany

[3] Department of Gynecology and Obstetrics, University Hospital, LMU Munich, Munich, Germany

[4] Division of General Pediatrics, Department of Pediatrics, Dr. von Hauner Children's Hospital, University Hospital, LMU Munich, Munich, Germany

[5] Division of Pediatric Haematology & Oncology, Department of Pediatrics, Dr. von Hauner Children's Hospital, University Hospital, LMU Munich, Munich, Germany

[6] Division of Pediatric Rheumatology, Department of Pediatrics, Dr. von Hauner Children's Hospital, University Hospital, LMU Munich, Munich, Germany

[7] Division of Pediatric Pulmonology, Department of Pediatrics, Dr. von Hauner Children's Hospital, University Hospital, LMU Munich, Munich, Germany

[8] Division of Pediatric Gastroenterology, Department of Pediatrics, Dr. von Hauner Children's Hospital, University Hospital, LMU Munich, Munich, Germany

[9] Neonatal Intensive Care Unit, Department of Pediatrics, Dr. von Hauner Children's Hospital, University Hospital, LMU Munich, Munich, Germany

[10] Paediatric Intensive Care Unit, Department of Pediatrics, Dr. von Hauner Children's Hospital, University Hospital, LMU Munich, Munich, Germany

[11] Department of Pediatric Cardiology and Pediatric Intensive Care, University Hospital, LMU Munich, Germany

**London School of Hygiene and Tropical Medicine (LSHTM)**

Principal Investigator: Shunmay Yeung [1,2,3]

Research group:

Manuel Dewez[1] David Bath[3], Elizabeth Fitchett[1], Fiona Cresswell[1]

1. Clinical Research Department, Faculty of Infectious and Tropical Disease, London School of Hygiene and Tropical Medicine, London
2. Department of Paediatrics, St. Mary's Imperial College Hospital, London
3. Department of Global Health and Development, Faculty of Public Health and Policy, London School of Hygiene and Tropical Medicine, London

**The UK DIAMONDS Clinical Network Collaborators are as follows:** Michael Carter, Paul Wellman, Shane Tibby, Jonathan Cohen, Francesca Davis, Julia Kenny, Marie White, Matthew Fish, Aislinn Jennings, Manu Shankar-Hari, Katy Fidler, Dan Agranoff, Vivien Richmond, Mathhew Seal, Saul Faust, Dan Owen, Ruth Ensom, Sarah McKay, Diana Mondo Mariya Shaji, Rachel Schranz, Prita Rughani, Amutha Anpananthar, Susan Liebeschuetz, Anna Riddell, Divya Divakaran, Louise Han, Nosheen Khalid, Ivone Lancoma-Malcolm, Jessica Schofield, Teresa Simagan, Mark Peters, Alasdair Bamford, Lauran O'Neill, Nazima Pathan, Esther Daubney, Deborah White, Melissa Heightman, Sarah Eisen, Terry Segal, Lucy

Wellings, Simon B Drysdale, Nicole Branch, Lisa Hamzah, Heather Jarman, Maggie Nyirenda, Lisa Capozzi, Emma Gardiner, Robert Moots, Magda Nasher, Anita Hanson, Michelle Linforth, Sean O'Riordan, Donna Ellis, Akash Deep, Ivan Caro, Fiona Shackley,, Arianna Bellini, Stuart Gormley, Samira Neshat, Barnaby J Scholefield, Ceri Robbins, Helen Winmill

#### 1.2. PERFORM Consortium

https://www.perform2020.org/

**PARTNER: IMPERIAL COLLEGE (UK)**

Chief investigator/PERFORM coordinator:

Michael Levin

Principal and co-investigators; work package leads (alphabetical order)

Aubrey Cunnington (grant application)

Tisham De (work package lead)

Jethro Herberg (Principle Investigator, Deputy Coordinator, grant application)

Myrsini Kaforou (grant application, work package lead)

Victoria Wright (grant application, Scientific Coordinator)

Research Group (alphabetical order)

Lucas Baumard; Evangelos Bellos; Giselle D'Souza; Rachel Galassini; Dominic Habgood-Coote; Shea Hamilton; Clive Hoggart; Sara Hourmat; Heather Jackson; Ian Maconochie; Stephanie Menikou; Naomi Lin; Samuel Nichols; Ruud Nijman; Ivonne Pena Paz; Oliver Powell, Priyen Shah; Ching-Fen Shen; Clare Wilson

Clinical recruitment at Imperial College Healthcare NHS Trust (alphabetical order)

Amina Abdulla; Ladan Ali; Sarah Darnell; Rikke Jorgensen; Sobia Mustafa; Salina Persand

Imperial College Faculty of Engineering

Molly Stevens (co-investigator), Eunjung Kim (research group); Benjamin Pierce (research group)

Clinical recruitment at Brighton and Sussex University Hospitals

Katy Fidler (Principle Investigator)

Julia Dudley (Clinical Research Registrar)

Research nurses: Vivien Richmond, Emma Tavliavini

Clinical recruitment at National Cheng Kung University Hospital

Ching-Fen Shen (Principal Investigator); Ching-Chuan Liu (Co-investigator); Shih-Min Wang (Co-investigator), funded by the Center of Clinical Medicine Research, National Cheng Kung University

**PARTNER: SERGAS (Spain)**

Principal Investigators

Federico Martinón-Torres[1]

Antonio Salas[1,2]

Research Group (alphabetical order)

Fernando Álvez González[1], Cristina Balo Farto[1], Ruth Barral-Arca[1,2], María Barreiro Castro[1], Xabier Bello[1,2], Mirian Ben García[1], Sandra Carnota[1], Miriam Cebey-López[1], María José Curras-Tuala[1,2], Carlos Durán Suárez[1], Luisa García Vicente[1], Alberto Gómez-Carballa[1,2], Jose Gómez Rial[1], Pilar Leboráns Iglesias[1], Federico Martinón-Torres[1], Nazareth Martinón-Torres[1], José María Martinón Sánchez[1], Belén Mosquera Pérez[1], Jacobo Pardo-Seco[1,2], Lidia Piñeiro Rodríguez[1], Sara Pischedda[1,2], Sara Rey Vázquez[1], Irene Rivero Calle[1], Carmen Rodríguez-Tenreiro[1], Lorenzo Redondo-Collazo[1], Miguel Sadiki Ora[1], Antonio Salas[1,2], Sonia Serén Fernández[1], Cristina Serén Trasorras[1], Marisol Vilas Iglesias[1].

[1] Translational Pediatrics and Infectious Diseases, Pediatrics Department, Hospital Clínico Universitario de Santiago, Santiago de Compostela, Spain, and GENVIP Research Group (www.genvip.org), Instituto de Investigación Sanitaria de Santiago, Universidad de Santiago de Compostela, Galicia, Spain.

[2] Unidade de Xenética, Departamento de Anatomía Patolóxica e Ciencias Forenses, Instituto de Ciencias Forenses, Facultade de Medicina, Universidade de Santiago de Compostela, and GenPop Research Group, Instituto de Investigaciones Sanitarias (IDIS), Hospital Clínico Universitario de Santiago, Galicia, Spain

[3] Fundación Pública Galega de Medicina Xenómica, Servizo Galego de Saúde (SERGAS), Instituto de Investigaciones Sanitarias (IDIS), and Grupo de Medicina Xenómica, Centro de Investigación Biomédica en Red de Enfermedades Raras (CIBERER), Universidade de Santiago de Compostela (USC), Santiago de Compostela, Spain

**PARTNER: RSU (Latvia)**

Principal Investigator

Dace Zavadska[1,2]

Other RSU group authors (in alphabetical order):

Anda Balode[1,2], Arta Bārzdiņa[1,2], Dārta Deksne[1,2], Dace Gardovska[1,2], Dagne Grāvele[2], Ilze Grope[1,2], Anija Meiere[1,2], Ieva Nokalna[1,2], Jana Pavāre[1,2], Zanda Pučuka[1,2], Katrīna Selecka[1,2], Aleksandra Sidorova[1,2], Dace Svile[2], Urzula Nora Urbāne[1,2].

[1] Riga Stradins university, Riga, Latvia.

[2] Children clinical university hospital, Riga, Latvia.

**PARTNER: Medical Research Council Unit The Gambia (MRCG) at LSHTM**

Principal Investigator

Effua Usuf

Additional Investigators

Kalifa Bojang

Syed M. A. Zaman

Fatou Secka

Suzanne Anderson

Anna RocaIsatou Sarr

Momodou Saidykhan

Saffiatou Darboe

Samba Ceesay

Umberto D'alessandro

Medical Research Council Unit The Gambia at LSHTM

P O Box 273,

Fajara, The Gambia

**PARTNER: ERASMUS MC-Sophia Children's Hospital (Netherlands**

Principal Investigator

Henriëtte A. Moll[1]

Research Group (alphabetical order)

Dorine M. Borensztajn[1], Nienke N. Hagedoorn, Chantal Tan [1], [1], Clementien L. Vermont[2], Joany Zachariasse [1]

Additional investigator

W Dik [3]

[1] Erasmus MC-Sophia Children's Hospital, Department of General Paediatrics, Rotterdam, the Netherlands

[2] Erasmus MC-Sophia Children's Hospital, Department of Paediatric Infectious Diseases & Immunology, Rotterdam, the Netherlands

[3] Erasmus MC, Department of immunology, Rotterdam, the Netherlands

**PARTNER: Swiss Pediatric Sepsis Study (Switzerland)**

Principal Investigators:

Philipp Agyeman, MD [1] (ORCID 0000-0002-8339-5444), Luregn J Schlapbach, MD, FCICM [2,3] (ORCID 0000-0003-2281-2598)

Clinical recruitment at University Children's Hospital Bern for PERFORM:

Christoph Aebi [1], Verena Wyss [1], Mariama Usman [1]

Principal and co-investigators for the Swiss Pediatric Sepsis Study:

Philipp Agyeman, MD [1], Luregn J Schlapbach, MD, FCICM [2,3], Eric Giannoni, MD [4,5], Martin Stocker, MD [6], Klara M Posfay-Barbe, MD [7], Ulrich Heininger, MD [8], Sara Bernhard-Stirnemann, MD [9], Anita Niederer-Loher, MD [10], Christian Kahlert, MD [10], Giancarlo Natalucci, MD [11], Christa Relly, MD [12], Thomas Riedel, MD [13], Christoph Aebi, MD [1], Christoph Berger, MD [12] for the Swiss Pediatric Sepsis Study

[1] Department of Pediatrics, Inselspital, Bern University Hospital, University of Bern, Switzerland

[2] Neonatal and Pediatric Intensive Care Unit, Children's Research Center, University Children's Hospital Zurich, University of Zurich, Zurich, Switzerland

[3]Child Health Research Centre, University of Queensland, and Queensland Children`s Hospital, Brisbane, Australia

[4] Clinic of Neonatology, Department Mother-Woman-Child, Lausanne University Hospital and University of Lausanne, Switzerland

[5] Infectious Diseases Service, Department of Medicine, Lausanne University Hospital and University of Lausanne, Switzerland

[6] Department of Pediatrics, Children's Hospital Lucerne, Lucerne, Switzerland

[7] Pediatric Infectious Diseases Unit, Children's Hospital of Geneva, University Hospitals of Geneva, Geneva, Switzerland

[8] Infectious Diseases and Vaccinology, University of Basel Children's Hospital, Basel, Switzerland

[9] Children's Hospital Aarau, Aarau, Switzerland

[10] Division of Infectious Diseases and Hospital Epidemiology, Children's Hospital of Eastern Switzerland St. Gallen, St. Gallen, Switzerland

[11] Department of Neonatology, University Hospital Zurich, Zurich, Switzerland

[12] Division of Infectious Diseases and Hospital Epidemiology, and Children's Research Center, University Children's Hospital Zurich, Switzerland

[13] Children's Hospital Chur, Chur, Switzerland

**PARTNER: Liverpool (UK)**

Principal Investigators

Enitan D Carrol[1,2,3]

Stéphane Paulus [1,]

Research Group (alphabetical order)

Elizabeth Cocklin[1], Rebecca Jennings[4], Joanne Johnston[4], Simon Leigh[1], Karen Newall[4], Sam Romaine[1]

[1] Department of Clinical Infection, Microbiology and Immunology, University of Liverpool Institute of Infection and Global Health , Liverpool, England

[2] Alder Hey Children's Hospital, Department of Infectious Diseases, Eaton Road, Liverpool, L12 2AP

[3] Liverpool Health Partners, 1st Floor, Liverpool Science Park, 131 Mount Pleasant, Liverpool, L3 5TF

[4]Alder Hey Children's Hospital, Clinical Research Business Unit, Eaton Road, Liverpool, L12 2AP

**PARTNER: NKUA (Greece)**

Principal investigator

Professor Maria Tsolia (all activities)

Investigator/Research fellow

Irini Eleftheriou (all activities)

Additional investigators

Recruitment: Maria Tambouratzi

Lab: Antonis Marmarinos (Quality Manager)

Lab: Marietta Xagorari

Kelly Syggelou

2nd Department of Pediatrics, National and Kapodistrian University of Athens,

"P. and A. Kyriakou" Children's Hospital

Thivon and Levadias

Goudi, Athens

**PARTNER : Micropathology Ltd (UK)**

Principal Investigator

Professor Colin Fink[1], Clinical Microbiologist

Additional investigators

Dr Marie Voice[1], Post doc scientist

Dr. Leo Calvo-Bado[1], Post doc scientist

[1] Micropathology Ltd, The Venture Center, University of Warwick Science Park, Sir William Lyons Road, Coventry, CV4 7EZ.

**PARTNER : Medical University of Graz (MUG, Austria)**

Principal Investigator

Werner Zenz[1] (all activities)

Co-investigators (alphabetical order)

Benno Kohlmaier[1] (all activities)

Nina A. Schweintzger[1] (all activities)

Manfred G. Sagmeister[1] (study design, consortium wide sample management)

Research team

Daniela S. Kohlfürst[1] (study design)

Christoph Zurl[1] (BIVA PIC)

Alexander Binder[1] (grant application)

Recruitment team, data managers, (alphabetical order)

Susanne Hösele[1], Manuel Leitner[1], Lena Pölz[1], Glorija Rajic[1],

Clinical recruitment partners (alphabetical order)

Sebastian Bauchinger[1], Hinrich Baumgart[4], Martin Benesch[3], Astrid Ceolotto[1], Ernst Eber[2], Siegfried Gallistl[1], Gunther Gores[5], Harald Haidl[1], Almuthe Hauer[1], Christa Hude[1], Markus Keldorfer[5], Larissa Krenn[4], Heidemarie Pilch[5], Andreas Pfleger[2], Klaus Pfurtscheller[4], Gudrun Nordberg[5], Tobias Niedrist[8], Siegfried Rödl[4], Andrea Skrabl-Baumgartner[1], Matthias Sperl[7], Laura Stampfer[5], Volker Strenger[3], Holger Till[6], Andreas Trobisch[5], Sabine Löffler[5]

[1] Department of Pediatrics and Adolescent Medicine, Division of General Pediatrics, Medical University of Graz, Graz, Austria

[2]Department of Pediatric Pulmonology, Medical University of Graz, Graz, Austria

[3]Department of Pediatric Hematooncoloy, Medical University of Graz, Graz, Austria

[4]Paediatric Intensive Care Unit, Medical University of Graz, Graz, Austria

[5]University Clinic of Paediatrics and Adolescent Medicine Graz, Medical University Graz, Graz,Austria

[6]Department of Paediatric and Adolescence Surgery, Medical University Graz, Graz, Austria

[7]Department of Pediatric Orthopedics, Medical University Graz, Graz, Austria

[8]Clinical Institute of Medical and Chemical Laboratory Diagnostics, Medical University Graz, Graz, Austria

**PARTNER: London School of Hygiene and Tropical Medicine (UK)**

WP 1 WP2, WP5

Principal Investigator:

Dr Shunmay Yeung[1,2 3] PhD, MBBS, FRCPCH, MRCP, DTM&H

Research Group

Dr Juan Emmanuel Dewez[1] MD, DTM&H, MSc

Prof Martin Hibberd [1] BSc, PhD

Mr David Bath[2] MSc, MappFin, BA(Hons)

Dr Alec Miners[2] BA(Hons), MSc, PhD

Dr Ruud Nijman[3] PhD MSc MD MRCPCH

Dr Catherine Wedderburn[1] BA, MBChB, DTM&H, MSc, MRCPCH

Ms Anne Meierford[1] MSc, BmedSc, BMBS

Dr Baptiste Leurent[4], PhD, MSc

1. Faculty of Infectious and Tropical Disease, London School of Hygiene and Tropical Medicine, London, UK
2. Faculty of Public Health and Policy, London School of Hygiene and Tropical Medicine, London, UK
3. Department of Paediatrics, St. Mary's Hospital Imperial College Hospital, London, UK
4. Faculty of Epidemiology and Population Health, London School of Hygiene and Tropical Medicine, London, UK

**PARTNER: Radboud University Medical Center (RUMC, Netherlands)**

Principal Investigators

Ronald de Groot [1], Michiel van der Flier [1,2,3], Marien I. de Jonge[1]

Co-investigators Radboud University Medical Center (alphabetical order)

Koen van Aerde[1,2], Wynand Alkema[1], Bryan van den Broek[1], Jolein Gloerich[1], Alain J. van Gool[1], Stefanie Henriet[1,2], Martijn Huijnen[1], Ria Philipsen[1], Esther Willems[1]

Investigators PeDBIG PERFORM DUTCH CLINICAL NETWORK (alphabetical order)

G.P.J.M. Gerrits[8], M. van Leur[8,] J. Heidema [4],L. de Haan[1,2] C.J. Miedema [5], C. Neeleman [1] C.C. Obihara [6], G.A. Tramper-Stranders7[6]

1. Radboud University Medical Center, Nijmegen, The Netherlands
2. Amalia Children's Hospital, Nijmegen, The Netherlands
3. Wilhelmina Children's Hospital, University Medical Center Utrecht, Utrecht, The Netherlands
4. St. Antonius Hospital, Nieuwegein, The Netherlands
5. Catharina Hospital, Eindhoven, The Netherlands
6. ETZ Elisabeth, Tilburg, The Netherlands
7. Franciscus Gasthuis, Rotterdam, The Netherlands
8. Canisius Wilhelmina Hospital, Nijmegen, The Netherlands

**PARTNER: Oxford (UK)**

Principal Investigators

Andrew J. Pollard[1,2], Rama Kandasamy[1,2], Stéphane Paulus [1,2]

Additional Investigators

Michael J. Carter[1,2], Daniel O'Connor[1,2], Sagida Bibi[1,2], Dominic F. Kelly[1,2], Meeru Gurung[3], Stephen Thorson[3], Imran Ansari[3], David R. Murdoch[4], Shrijana Shrestha[3].

[1]Oxford Vaccine Group, Department of Paediatrics, University of Oxford, Oxford, United Kingdom.

[2]NIHR Oxford Biomedical Research Centre, Oxford, United Kingdom.

[3]Paediatric Research Unit, Patan Academy of Health Sciences, Kathmandu, Nepal.

[4]Department of Pathology, University of Otago, Christchurch, New Zealand.

**PARTNER: Newcastle University, Newcastle upon Tyne, (UK)**

Principal Investigator

Marieke Emonts [1,2,3] (all activities)

Co-investigators

Emma Lim[2,3,7] (all activities)

Lucille Valentine[4]

Recruitment team (alphabetical), data-managers, and GNCH Research unit

Karen Allen[5], Kathryn Bell[5], Adora Chan[5], Stephen Crulley[5], Kirsty Devine[5], Daniel Fabian[5], Sharon King[5], Paul McAlinden[5], Sam McDonald[5], Anne McDonnell2,[5], Ailsa Pickering[2,5], Evelyn Thomson[5], Amanda Wood[5], Diane Wallia[5], Phil Woodsford[5],

Sample processing: Frances Baxter[5], Ashley Bell[5], Mathew Rhodes[5]

PICU recruitment

Rachel Agbeko[8]

Christine Mackerness[8]

Students MOFICHE

Bryan Baas[2], Lieke Kloosterhuis[2], Wilma Oosthoek[2]

Students/medical staff PERFORM

Tasnim Arif[6], Joshua Bennet[2], Kalvin Collings[2], Ilona van der Giessen[2], Alex Martin[2], Aqeela Rashid[6], Emily Rowlands[2], Gabriella de Vries[2], Fabian van der Velden[2]

Engagement work/ethics/cost effectiveness

Lucille Valentine [4], Mike Martin[9], Ravi Mistry[2], Lucille Valentine[4]

[1] Translational and Clinical Research Institute, Newcastle University, Newcastle upon Tyne UK

[2]Great North Children's Hospital, Paediatric Immunology, Infectious Diseases & Allergy, Newcastle upon Tyne Hospitals NHS Foundation Trust, Newcastle upon Tyne, United Kingdom.

[3]NIHR Newcastle Biomedical Research Centre based at Newcastle upon Tyne Hospitals NHS Trust and Newcastle University, Westgate Rd, Newcastle upon Tyne NE4 5PL, United Kingdom

[4]Newcastle University Business School, Centre for Knowledge, Innovation, Technology and Enterprise (KITE), Newcastle upon Tyne, United Kingdom

[5]Great North Children's Hospital, Research Unit, Newcastle upon Tyne Hospitals NHS Foundation Trust, Newcastle upon Tyne, United Kingdom.

[6]Great North Children's Hospital, Paediatric Oncology, Newcastle upon Tyne Hospitals NHS Foundation Trust, Newcastle upon Tyne, United Kingdom.

[7]Population Health Sciences Institute, Newcastle University, Newcastle upon Tyne, UK

[8]Great North Children's Hospital, Paediatric Intensive Care Unit, Newcastle upon Tyne Hospitals NHS Foundation Trust, Newcastle upon Tyne, United Kingdom.

[9]Northumbria University, Newcastle upon Tyne, United Kingdom.

**PARTNER: LMU Munich (Germany)**

Principal Investigator

Ulrich von Both[1,2] MD, FRCPCH (all activities)

Research group

Laura Kolberg[1] MSc (all activities)

Manuela Zwerenz[1] MSc, Judith Buschbeck[1] PhD

Clinical recruitment partners (alphabetical order)

Christoph Bidlingmaier[3], Vera Binder[4], Katharina Danhauser[5], Nikolaus Haas[10], Matthias Griese[6], Tobias Feuchtinger[4], Julia Keil[9], Matthias Kappler[6], Eberhard Lurz[7], Georg Muench[8], Karl Reiter[9], Carola Schoen[9]

[1]Div. Paediatric Infectious Diseases, Hauner Children's Hospital, University Hospital, Ludwig Maximilians University (LMU), Munich, Germany

[2]German Center for Infection Research (DZIF), Partner Site Munich, Munich, Germany

[3]Div. of General Paediatrics, [4]Div. Paediatric Haematology & Oncology, [5]Div. of Paediatric Rheumatology, [6]Div. of Paediatric Pulmonology, [7]Div. of Paediatric Gastroenterology, [8]Neonatal Intensive Care Unit, [9]Paediatric Intensive Care Unit Hauner Children's Hospital, University Hospital, Ludwig Maximilians University (LMU), Munich, Germany, [10]Department Pediatric Cardiology and Pediatric Intensive Care, University Hospital, Ludwig Maximilians University (LMU), Munich, Germany

**PARTNER: bioMérieux (France)**

Principal Investigator

François Mallet[1,2, 3]

Research Group

Karen Brengel-Pesce[1,2, 3]

Alexandre Pachot[1]

Marine Mommert[1,2]

[1]Open Innovation & Partnerships (OIP), bioMérieux S.A., Marcy l'Etoile, France

[2] Joint research unit Hospice Civils de Lyon – bioMérieux, Centre Hospitalier Lyon Sud, 165 Chemin du Grand Revoyet, 69310 Pierre-Bénite, France

[3] EA 7426 Pathophysiology of Injury-induced Immunosuppression, University of Lyon1-Hospices Civils de Lyon-bioMérieux, Hôpital Edouard Herriot, 5 Place d'Arsonval, 69437 Lyon Cedex 3, France

**PARTNER: University Medical Centre Ljubljana (Slovenia)**

Principal Investigator

Marko Pokorn[1,2,3] MD, PhD

Research Group

Mojca Kolnik[1] MD, Katarina Vincek[1] MD, Tina Plankar Srovin[1] MD, PhD, Natalija Bahovec[1] MD, Petra Prunk[1] MD, Veronika Osterman[1] MD, Tanja Avramoska[1] MD

[1]Department of Infectious Diseases, University Medical Centre Ljubljana, Japljeva 2, SI-1525 Ljubljana, Slovenia

[2]University Childrens' Hospital, University Medical Centre Ljubljana, Ljubljana, Slovenia

[3]Department of Infectious Diseases and Epidemiology, Faculty of Medicine, University of Ljubljana, Slovenia

**PARTNER: Amsterdam, Academic Medical Hospital & Sanquin Research Institute (Netherlands)**

Principal Investigator

Taco Kuijpers [1,2]

Co-investigators

Ilse Jongerius [2]

Recruitment team (EUCLIDS, PERFORM)

J.M. van den Berg[1], D. Schonenberg[1], A.M. Barendregt[1], D. Pajkrt[1], M. van der Kuip[1,3], A.M. van Furth[1,3]

Students PERFORM

Evelien Sprenkeler [2], Judith Zandstra [2]

Technical support PERFORM

G. van Mierlo [2], J. Geissler [2]

[1] Amsterdam University Medical Center (Amsterdam UMC), location Academic Medical Center (AMC), Dept of Pediatric Immunology, Rheumatology and Infectious Diseases, University of Amsterdam, Amsterdam, the Netherlands

[2] Sanquin Research Institute, & Landsteiner Laboratory at the AMC, University of Amsterdam, Amsterdam, the Netherlands.

[3] Amsterdam University Medical Center (Amsterdam UMC), location Vrije Universiteit Medical Center (VUMC), Dept of Pediatric Infectious Diseases and Immunology, Free University (VU), Amsterdam, the Netherlands (former affiliation)

#### 1.3 UK Kawasaki Disease Genetic Consortium

Dept of Paediatrics, Imperial College London; Professor M Levin, Rachel Galassini , Dr Victoria Wright, Dr Jethro Herberg

Addenbrookes Hospital, Cambridge: Dr Y Singh (PI), J Bytham, J Sharp

Airedale General Hospital Dr P Bala (PI), A Kitching

Alder Hey children's Hospital: Dr S Paulus (PI), Prof E Carol (PI), Dr B Larru (PI). S Wadeson, J Johnstone, R Jennings

Birmingham Children's Hospital; Dr A Chickermane (PI), K Cotter

Bradford Royal Infirmary: Dr H Jepps (PI); T Booth, R Swingler

Bristol Royal Infirmary: Prof R Tulloh (PI); Karen Sheehan

Burton Hospital: Dr M Ahmed (PI), S Boswell, C Backhouse

Calderdale Royal Hospital Dr M Olabi (PI), KU Rahman (PI), S Kilroy, M Home

Durham & Darlington NHS Trust: Dr T Banerjee (PI), Dr G Nyamugunduru (PI), A Cowton, D Egginton

East Surrey Hospital; Dr M Jawad (PI)L Bailey

Evelina Children's Hospital: Dr E Menson (PI)

Great Ormond Street Hospital, Dr P Brogan (PI), Y Glackin

Harrogate Hospital: Miss C Brunskill (PI)

Heartlands Hospital Dr S Hackett (PI), J Daglish

Hereford County Hospital Dr S Meyrick (PI), E Collins

Hull University Teaching Hospital, Mr D Bolton (PI)

Imperial College Healthcare NHS Trust; Dr J Herberg (PI), S Gormley, S Mustafa

Ipswich Hospital: Dr P Desai (PI) L Hunt

Kingston Hospital Dr T Chawatama (PI), Dr S Luck (PI), J Crooks, T O'Brien

Leeds General Infirmary Dr S O'Riordan (PI), N Balatoni, N Maher

Macclesfield General Hospital: Dr Chandrasekaran (PI), N Keenan

New Cross Hospital, Wolverhampton, Dr K Davies (PI), S Kempson, C Busby

North Manchester General Hospital: Dr E Odeka (PI), G O'Connor

North Tees & Hartlepool Dr I Haar (PI), G Osborne, H Walker

Northwick Park Hospital: Dr A Williams (PI)

Oldham Hospital Dr E Odeka (PI), L Woodward, C Rishton

Peterborough City Hospital; Dr V Puthi (PI), A Pearson, P Goodyear

Pinderfields General Hospital, Dr C Davidson (PI), Dr N De Vere (PI), G Castle

Royal Albert Edward Infirmary, Dr M Farrier (PI), N Pemberton

Royal Bolton Hospital: Dr S Misra (PI), C Fish, P Graham, J Henry

Royal Lancaster Infirmary, Dr A Olabi (PI) K Allison

Royal Shrewsbury Hospital: Dr A Kannivelu (PI), Mr J Jones (PI)

Royal Stoke University Hospital: Dr J Alexander (PI) E Roe, R Pringle, A Cope

Sheffield Children's Hospital, Dr F Shackley (PI), S Gormley

South Tees Hospital: Dr R Kumar (PI), GMcilhinney, S Armstrong

St George's Hospital, Tooting, Prof P Heath (PI), E Vitale, J Stuart

St Richard's Hospital, Chichester: Dr N Brennan (PI), S Floyd

Stepping Hill Hospital, Stockport, Dr C Cooper, S Bennett

Tameside Hospital, Dr A Petkar (PI), Dr C Greenway (PI), W Hulse

The Royal Alexandra Hospital, Brighton; Dr K Fidler (PI), K Moscovici, S Sobowieck Kouman

The Royal Brompton Hospital, Dr F Franklin (PI), Dr M Bartsota

The Royal Cornwall Hospital, Dr N Venkata (PI), Dr A Prendiville (PI), Dr O Elmasry (PI), Mrs H Osborne (PI), G Craig, B Bromage

Torbay Hospital, Dr M Raman (PI), Ms P Fitzell (PI), H Bearne, J Palmer

UK Kawasaki Support Group S Davidson, N Clements
